# Reclassifying Ablation Energy Risk Profiles in Atrial Fibrillation A Narrative-Adjudicated Comparative Analysis of the MAUDE Database

**DOI:** 10.64898/2026.09.30.26364455

**Authors:** Anish S. Shah, Reese Fuller, Emile G. Daoud, Bobby Endo, Ashley Goodwin, Roberto Martinez-Parachini, Marc C. Engels, Immo Lehmann, Klitos Konstantinidis, Ravi Ranjan, T. Jared Bunch

## Abstract

**Background:** Pulsed field ablation (PFA) has been rapidly adopted for atrial fibrillation (AF) on the premise of greater safety than thermal ablation. Real-world signals of hemolysis, coronary spasm, and neurologic events have since emerged, and prior Manufacturer and User Facility Device Experience (MAUDE) analyses relied on term matching without prospective signal detection.

**Methods:** We queried MAUDE for AF ablation catheter reports received January 2016 through June 2026 and assigned each to PFA, radiofrequency ablation (RFA), or cryoballoon ablation. We adjudicated injury and death narratives into predefined complication families and severity tiers with a large language model, checked against a larger model and by physician review. We calculated reporting odds ratios (ROR) and Bayesian shrinkage estimators compared against an empirical null, applied interrupted time series and prospective sequential detection, and repeated estimates on a 2026 hold-out.

**Results:** Among 44,623 reports, 30.1% were PFA, 50.9% RFA, and 19.0% cryoballoon, with 91.8% model agreement on event occurrence (*κ* = 0.83). PFA carried an ROR of 527 (95% CI, 33, 8,509) for hemolysis, 2.83 (2.55, 3.16) for arrhythmia, 2.83 (2.30, 3.48) for coronary, and 2.39 (2.10, 2.73) for cerebrovascular events, against 0.024 (0.008, 0.075) for esophageal and 0.34 (0.32, 0.37) for pericardial events. The cerebrovascular excess was largest for VARIPULSE, but without it the class proportional reporting ratio was 1.36 (1.13, 1.65), concordant with the 1.56 measured in *EMBOL-AF*. In the hold-out, the cerebrovascular, arrhythmia and hemolysis signals replicated, and coronary events kept their direction without meeting the signal rule. Prospective detection flagged cerebrovascular events 13 months before *AVANT GUARD* published its neurologic events.

**Conclusions:** PFA showed disproportionate reporting of hemolysis, cerebrovascular, coronary, and arrhythmic events across sequential testing. Machine learning methods allowed for flagging these signals months before confirmatory trials, demonstrating a scalable tool for early detection of procedural risk.

**CLINICAL PERSPECTIVE:** **What is Known**

- Pulsed field ablation has been adopted rapidly for atrial fibrillation on the premise of fewer collateral injuries than thermal ablation.
- Hemolysis, coronary spasm, and neurologic events have since emerged in case series, registries, and a paused trial, while prior analyses of Food and Drug Administration device reports relied largely on term matching without narrative adjudication at scale.

**What the Study Adds**

- In injury and death reports adjudicated at the narrative level, pulsed field ablation was reported more often than thermal ablation for hemolysis, arrhythmia, coronary, and cerebrovascular events and less often for esophageal and pericardial injury.
- The cerebrovascular reporting excess was largest for the VARIPULSE catheter but persisted with that platform removed and concentrated in minor stroke and systemic embolism, suggesting an embolic signal not confined to one platform.
- Sequential detection run on reports adjudicated by a large language model would have flagged the cerebrovascular signal 13 months before *AVANT GUARD* published its neurologic events, suggesting a role for automated surveillance of new ablation technologies.

## INTRODUCTION

Pulsed field ablation (**PFA**) for cardiac tissue uses selective waveforms to induce irreversible electroporation to ablate conducting myocardial tissue, rather than thermal ablative energy. In theory, this selectivity offers an improved safety profile over thermal ablation technology.^1^ The first generation of catheters are now broadly available after a rapid sequence of United States approvals, with PulseSelect (Medtronic) in December 2023, FARAPULSE (Boston Scientific) in January 2024, Affera Sphere-9 (Medtronic) in October 2024, VARIPULSE (Biosense Webster) in November 2024, and Volt (Abbott) in December 2025. Large post-approval registries have reproduced the efficacy of pulmonary vein isolation for atrial fibrillation (**AF**) seen in the original pivotal trials and have reported a safety and efficacy over that of radiofrequency ablation (**RFA**) and cryoballoon ablation (**CA**), with no esophageal injury and no persistent phrenic nerve palsy.^2–4^ *MANIFEST-US* has since extended that experience with the pentaspline catheter to over 40,000 patients.^5^

The reported safety profile rests on data that differs from clinical experience in several critical ways. The largest of these registries are associated with the manufacturer, with site selection, event evaluation, and adjudication organized around the catheter under the respective study.^2–4^ Similarly, the pivotal trials are appropriately sponsored and enroll selected patients with experienced operators, and are generally powered for composite safety and efficacy endpoints.^6,7^ Composite outcomes can bias signal away or towards the null depending on how endpoints are specified, as seen in non-inferiority trials.^8^ Because these trials often represent a best case scenario, the real-world experience can unearth complications that are under-represented and identify the crucial need for prospective signal detection to help respond to risks and patterns that can emerge at scale.

Specific risks and event types have surfaced in smaller series and single-platform evaluations with PFA, the most common being hemolysis with subsequent kidney injury^9–13^, and coronary spasm following energy application proximal to the coronary arteries.^14–16^ These risks have been assessed and confirmed prospectively as well in *NEMESIS-PFA*, which found myocardial injury, hemolysis, and renal dysfunction more prominent than with RFA.^17^ Neurologic events have instead been more prominent in the regulatory records, the most well known of which is the increased stroke risk seen with VARIPULSE, leading to a pause in January 2025 with subsequent irrigation instructions for use (**IFU**) corrections that summer.^18^ Similarly, in the *AVANT GUARD* trial, there was a pause in enrollment in October 2024 after an early increase in neurologic events in the FARAPULSE arm, subsequently reported upon publication in April 2026.^19^ The VARIPULSE signal was replicated in a real-world assessment of events in the Food and Drug Administration (**FDA**) Manufacturer and User Facility Device Experience (**MAUDE**) database.^18,20^ The increased embolic risk has subsequently started to be noted in other registries and single-center studies.^21,22^

The delay between real-world data signals and publication data leads to a critical gap in safety monitoring, such as when the Sprint Fidelis lead fracture signal was identified a year before its recall.^23^ The purported solution, such as the MAUDE database, however remains limited by the lack of a public denominator, under-reporting, lack of event ascertainment and adjudication, and the idiosyncratic issues that may arise that are platform-specific. The sheer volume of MAUDE exceeds what can be read by hand, term matching of complication codes is insufficient to identify underlying signals outside of smaller, less common procedure types,^24^ and understanding when and which signals warrant intervention in a noisy baseline are both difficult to perform and interpret. However, prospective signal detection analyses in MAUDE should not be abandoned. In order to identify potential early signs of complications, and allow for more rapid shifts in clinical practice when appropriate, we must have a thorough and thoughtful approach in how to evaluate data in a prospective manner.^18^

In this study, we sought to place the real-world experience of complications with AF ablation in context of the shifting modalities of performing AF ablation over the past decade. We utilized the MAUDE database to not only demonstrate differences and disproportionality in reporting patterns, but also assess the importance of prospective signal detection methods in ways that are often under-recognized in the literature. Moreover, we sought to contextualize how ablation complications are also responsive to not only energy modality, but the force of external factors, from FDA advisories, preliminary data in clinical trials, and regulatory changes. We hypothesized that the risk profile with PFA is markedly different than with thermal ablation, and that the persistent pattern of complications warrant continued scrutiny when considering how to define acceptable shifting of risk in patient outcomes.

Prior MAUDE evaluations of PFA catalogued adverse events and largely attributed the neurologic signal to a single platform, but each was constrained by term matching without narrative confirmation, by early-adoption reporting windows that predated the current device landscape, and by the absence of a thermal comparator; limitations that risk both misclassifying event types and mistaking notoriety bias for a true class effect. These are not incremental gaps: they directly determine whether the reported neurologic and hemolytic signals are real, generalizable, and attributable to the modality itself, which is precisely the question clinicians and regulators now face and will face with rapidly evolving PFA technologies. A repeat analysis is therefore necessary to resolve them, and here we adjudicate every report at the narrative level and compare against both an empirical null and a ten-year thermal baseline to separate a genuine class effect from disproportionality artifacts. We further use MAUDE to test the under-recognized value of prospective signal detection quantifying how many months sooner routine surveillance could have flagged risks that trials and registries later confirmed, and to examine how complication reporting responds to external forces such as FDA advisories and revised IFU. We hypothesized that the PFA risk profile differs markedly from thermal ablation and that its persistent, platform-and tier-specific pattern warrants continued scrutiny when defining an acceptable shift in patient risk.

## METHODS

We queried the FDA MAUDE database for reports received from January 2016 through June 2026, dated by FDA receipt. Reports from 2015 and prior were excluded as patient problem terms were absent in the preceding years, limiting an equivalent adjudication. We searched five product codes that encompassed the catheters used for AF ablation (QZI, OAE, LPB, OAD, and DRA). Every report carries an FDA event type, which was split into injury or death reports, in which a patient outcome is recorded, and malfunction reports, which describe a device issue, and may not have a patient outcome. Injury or death reports were the basis for all clinical estimands, as malfunction reports do not regularly contain a clinical outcome, and malfunction reports were the basis for all device event estimands. The workflow is shown in **Figure 1**.

**Figure 1:**
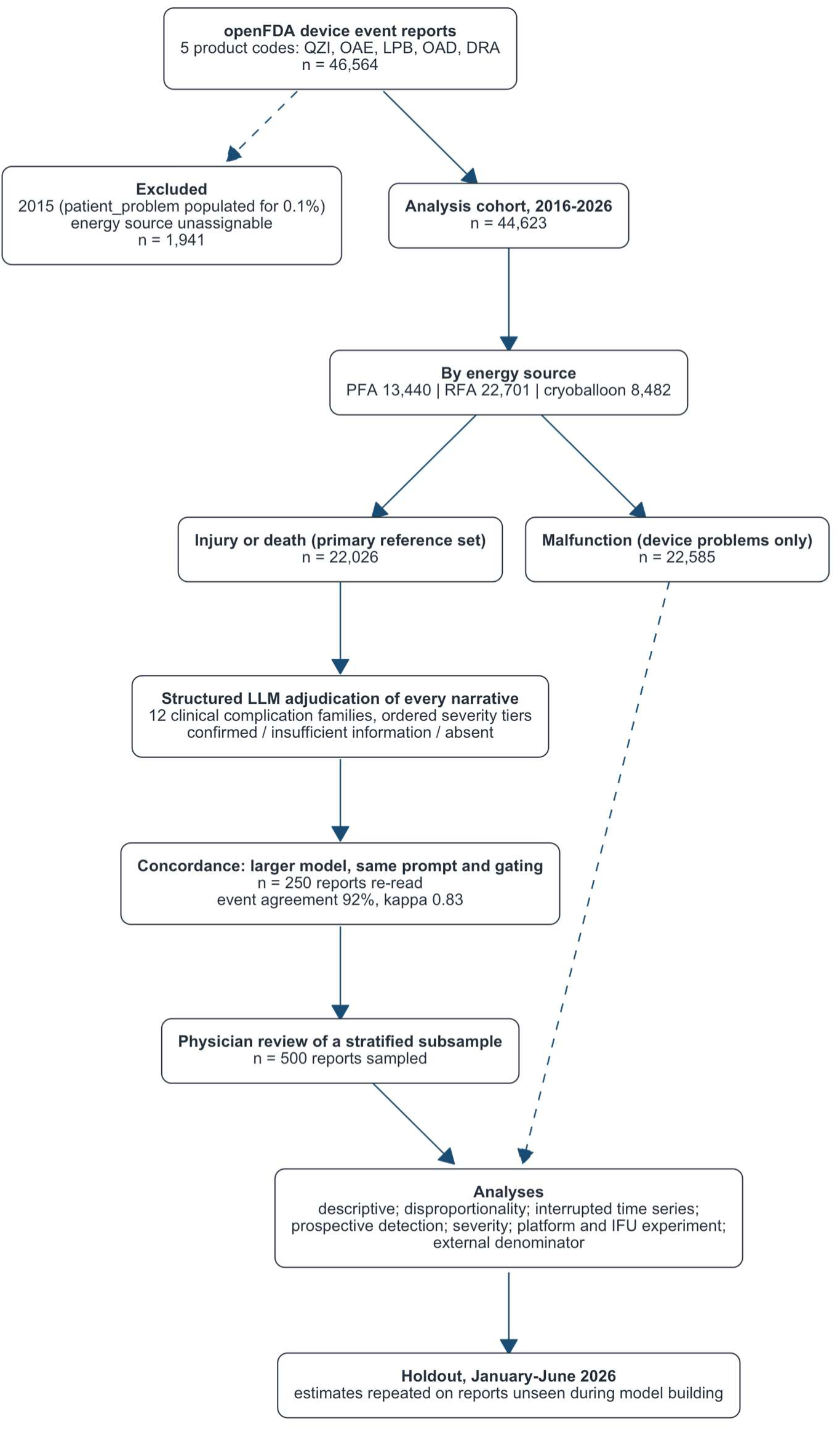
Report selection, machine adjudication, and analysis. Counts are reports, not patients or procedures. The Injury or Death stream is the primary reference set for every clinical estimate; malfunction reports carry a patient outcome too rarely to serve as one and are analysed only for device problems. Adjudication was performed by a structured-output model over every narrative, re-read on the same prompt and gating by a larger model, with a physician-reviewed subsample.

We assigned each report to PFA, RFA, or CA by product code, and compared across all three energy modalities. PFA reports were assigned a platform, restricted to the four with the largest market share: FARAPULSE (Boston Scientific), PulseSelect (Medtronic), VARIPULSE (Biosense Webster), and Affera Sphere-9 (Medtronic). When the code could not distinguish between the energy source or platform, we manually reviewed the report to identify the brand. MAUDE codes each report with International Medical Device Regulators Forum terms chosen by the reporter at submission, either patient problem terms for injury and death, or device problem terms for malfunctions.

The clinical content of a report is comprised of the patient problem term and the associated narrative text, entered by the manufacturer or by a voluntary reporter such as a facility or a physician. We defined complication families from the definitions used in consensus statements and the large PFA registries,^3,25,26^ and subdivided each into severity tiers, as seen in **Table S1**. We then performed event adjudication using both the specified problem terms and the narrative text, with structured instructions and complication definitions. The problem terms were used to select potential candidate complication families, and a large language model (**LLM**), GPT-5 Nano (OpenAI), read the narrative once for each candidate family and returned a structured answer for every complication tier, with an insufficient information outcome that did not count towards the event count. The report term that was selected at submission was only counted when the narrative confirmed it, which differentiates this analysis from keyword or regular expression matching approaches. We measured the reliability of the LLM findings with a larger model, GPT-5.6 Luna (OpenAI), which performed an adversarial adjudication of a stratified sample of reports. We report the raw agreement and Cohen’s kappa on whether an event occurred, and a physician reviewed the sample.

Our statistical approach was based on additive signal detection strategies. The exposure was energy source and the outcome was a confirmed complication family. Every estimate describes reporting rather than incidence, as MAUDE provides no procedural denominator. We report descriptive counts and proportions by energy source and year. We assessed disproportionality overall, by year, and by quarter from the 2 × 2 table of each energy source against all others, as the reporting odds ratio (**ROR**) and the proportional reporting ratio (**PRR**).^27^ For sparse cells, we performed the Haldane and Anscombe correction by adding 0.5 to every cell, to avoid undefined zero cells.^28,29^ A signal required a PRR ≥ 2 with *χ*^2^ ≥ 4 and at least 3 events, or an ROR lower confidence bound above 1. We added two shrinkage estimators built for sparse cells, the information component and the empirical Bayes geometric mean (**EBGM**).^30,31^ A shrinkage signal required the empirical Bayes 5th percentile (**EB05**) to be at least 2. We applied the Benjamini-Hochberg procedure across the Fisher exact P-values to correct for false discovery rates over different time periods assessed.^32^ At baseline, we assessed the thermal complication associations with the established injury rates (e.g. phrenic nerve injury with CA, or esophageal injury with RFA), and paired those against where PFA was expected to be protective, as the expected null. The estimates over the null pairs gave an empirical null, to assess the reporting gap size between comparators.^18^

We used interrupted time series as a surrogate for a change in signal, fitting segmented regressions of monthly complication counts, offset for the number of reports, so the modeled quantity was the share of reports carrying the complication, both for the pooled series and for PFA against the concurrent thermal series as a control.^33^ The change point was set at January 2024, the first full month after FDA approval of PFA, with the predicted slope trend prior as the counterfactual.

We additionally ran a surveillance program in a prospective and retrospective manner. Prospectively, we used a Shewhart chart with Laney’s correction for overdispersion, tracking monthly reporting against a 2022 – 2023 baseline,^34^ a Poisson cumulative sum (**CUSUM**) chart, and a maximized sequential probability ratio test (**MaxSPRT**) to compare simultaneous PFA events with thermal reporting events.^35,36^ A retrospective scan located a change point for each time series by identifying segmented fits with minimal deviance. All estimates were compared with dates of approvals, safety actions, reporting policy changes, and large clinical studies.

We utilized two methods to assess the impact of a synthetic external denominator to estimate the potential risk. We first inverted the disproportionality test (reverse binomial test) by quarter to find the smallest share that PFA would have to account for from all theoretical AF ablations that would allow the PFA share of complication reports to be unsurprising, under a binomial test at *α* = 0.05. The second assumed no under-reporting of PFA events, and treated PFA reports as the numerator over the published rates of complications. That allowed us to hypothesize a theoretical denominator of procedures that would be required to produce the observed number of reports, segregated by the rates available from recent large studies: MANIFEST-17K,^3^ MANIFEST-PF,^2^ EU-PORIA,^4^ EMBOL-AF,^22^ and NEMESIS-PFA.^17^ EMBOL-AF served as the calibration anchor because it measured PFA and thermal events in the same centers over the same years with a denominator.

As a sensitivity analysis, we examined each complication family on its own, focusing on the most common complications of cerebrovascular, coronary, pericardial, and arrhythmia events. We additionally examined complications by individual PFA platform and in a leave-one-platform-out analysis that removed each platform in turn from the class estimate, allowing us to assess the effect of FDA advisories and updates to the IFU on reporting rates as a natural experiment.^18^ Specifically, we examined the VARIPULSE platform, which had an FDA advisory in January 2025 with subsequent June 2025 updates to the IFU for irrigation flow, and FARAPULSE against the neurological event rates found in the *AVANT GUARD* trial to assess whether the signal was detectable prior to the study’s publication.^19^ We additionally re-estimated the disproportionality on reports received from January through June 2026 alone, against estimates from reports received through December 2025, as a later period comparison.

Statistical detail and software are in the Supplemental Material. Analyses performed with R version 4.6.1 (R Core Team, Vienna, Austria), and MAUDE querying and retrieval from openFDA were through the card package.^37,38^

## RESULTS

We identified 44,623 reports received from 2016 through 2026, of which 13,440 (30.1%) were PFA, 22,701 (50.9%) RFA, and 8,482 (19.0%) CA (**Figure 1**, **Table 1**). Within PFA, FARAPULSE accounted for 9,895 reports, PulseSelect for 1,937, Affera Sphere-9 for 928, and VARIPULSE for 491, with 189 reports on other PFA platforms. PFA reports were 34.9% of the reports received in 2024 and 67.3% of those received in 2025. The injury or death reports had 4,705 PFA, 13,153 RFA, and 4,168 CA reports.

**Table 1:**
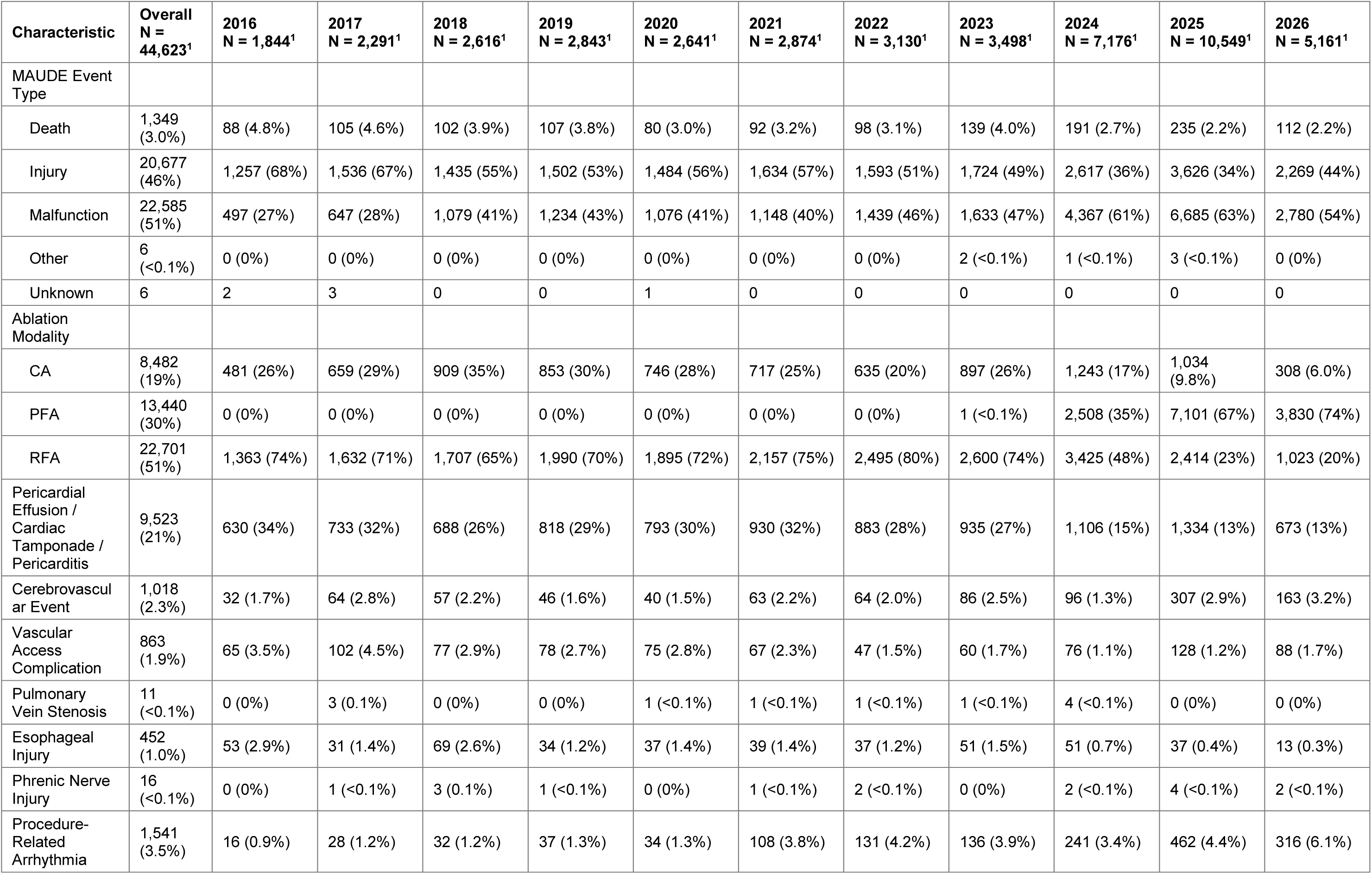
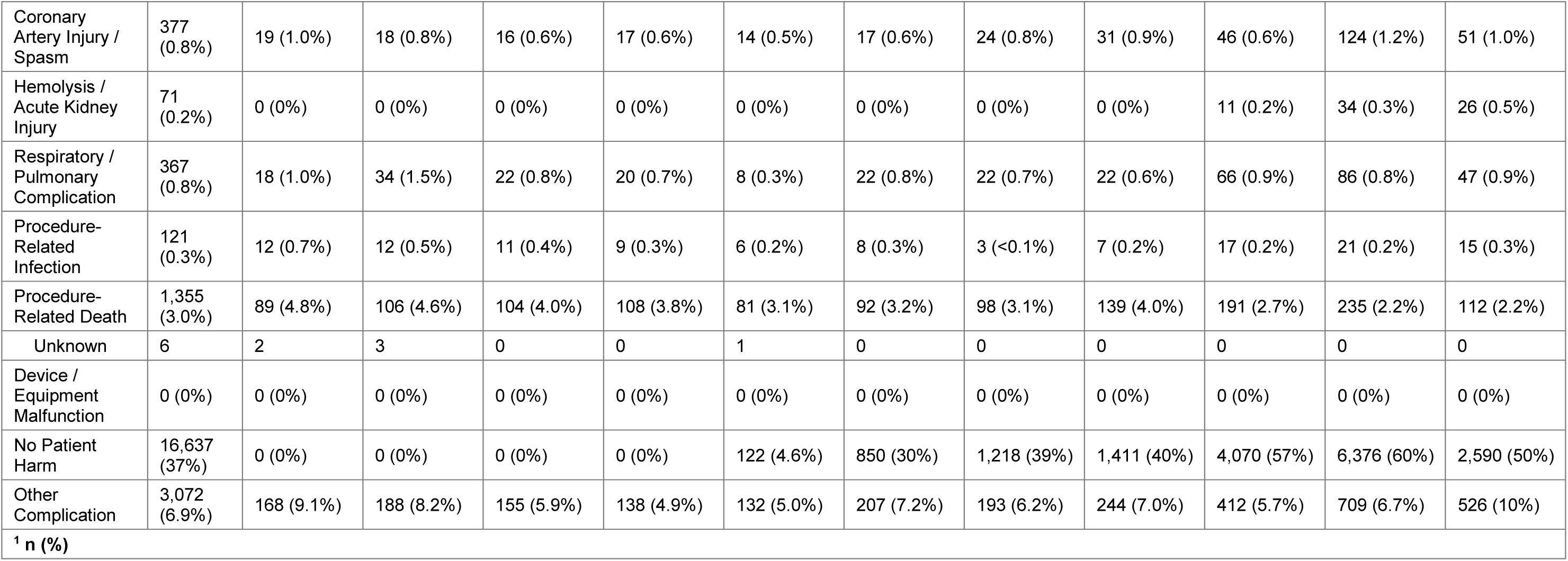
Composition of the adjudicated report set by year. Counts are reports, not patients or procedures, so no row is an incidence. Percentages are of that year’s column. The complication rows are confirmed adjudications and are not mutually exclusive, since one report can carry more than one family.

| Characteristic | Overall<br>N =<br>44,623<sup>1</sup> | 2016<br>N = 1,844<sup>1</sup> | 2017<br>N = 2,291<sup>1</sup> | 2018<br>N = 2,616<sup>1</sup> | 2019<br>N = 2,843<sup>1</sup> | 2020<br>N = 2,641<sup>1</sup> | 2021<br>N = 2,874<sup>1</sup> | 2022<br>N = 3,130<sup>1</sup> | 2023<br>N = 3,498<sup>1</sup> | 2024<br>N = 7,176<sup>1</sup> | 2025<br>N = 10,549<sup>1</sup> | 2026<br>N = 5,161<sup>1</sup> |
| --- | --- | --- | --- | --- | --- | --- | --- | --- | --- | --- | --- | --- |
| MAUDE Event Type |  |  |  |  |  |  |  |  |  |  |  |  |
| Death | 1,349<br>(3.0%) | 88 (4.8%) | 105 (4.6%) | 102 (3.9%) | 107 (3.8%) | 80 (3.0%) | 92 (3.2%) | 98 (3.1%) | 139 (4.0%) | 191 (2.7%) | 235 (2.2%) | 112 (2.2%) |
| Injury | 20,677<br>(46%) | 1,257 (68%) | 1,536 (67%) | 1,435 (55%) | 1,502 (53%) | 1,484 (56%) | 1,634 (57%) | 1,593 (51%) | 1,724 (49%) | 2,617 (36%) | 3,626 (34%) | 2,269 (44%) |
| Malfunction | 22,585<br>(51%) | 497 (27%) | 647 (28%) | 1,079 (41%) | 1,234 (43%) | 1,076 (41%) | 1,148 (40%) | 1,439 (46%) | 1,633 (47%) | 4,367 (61%) | 6,685 (63%) | 2,780 (54%) |
| Other | 6<br>($<0.1\%$) | 0 (0%) | 0 (0%) | 0 (0%) | 0 (0%) | 0 (0%) | 0 (0%) | 0 (0%) | 2 ($<0.1\%$) | 1 ($<0.1\%$) | 3 ($<0.1\%$) | 0 (0%) |
| Unknown | 6 | 2 | 3 | 0 | 0 | 1 | 0 | 0 | 0 | 0 | 0 | 0 |
| Ablation Modality |  |  |  |  |  |  |  |  |  |  |  |  |
| CA | 8,482<br>(19%) | 481 (26%) | 659 (29%) | 909 (35%) | 853 (30%) | 746 (28%) | 717 (25%) | 635 (20%) | 897 (26%) | 1,243 (17%) | 1,034<br>(9.8%) | 308 (6.0%) |
| PFA | 13,440<br>(30%) | 0 (0%) | 0 (0%) | 0 (0%) | 0 (0%) | 0 (0%) | 0 (0%) | 0 (0%) | 1 ($<0.1\%$) | 2,508 (35%) | 7,101 (67%) | 3,830 (74%) |
| RFA | 22,701<br>(51%) | 1,363 (74%) | 1,632 (71%) | 1,707 (65%) | 1,990 (70%) | 1,895 (72%) | 2,157 (75%) | 2,495 (80%) | 2,600 (74%) | 3,425 (48%) | 2,414 (23%) | 1,023 (20%) |
| Pericardial Effusion / Cardiac Tamponade / Pericarditis | 9,523<br>(21%) | 630 (34%) | 733 (32%) | 688 (26%) | 818 (29%) | 793 (30%) | 930 (32%) | 883 (28%) | 935 (27%) | 1,106 (15%) | 1,334 (13%) | 673 (13%) |
| Cerebrovascular Event | 1,018<br>(2.3%) | 32 (1.7%) | 64 (2.8%) | 57 (2.2%) | 46 (1.6%) | 40 (1.5%) | 63 (2.2%) | 64 (2.0%) | 86 (2.5%) | 96 (1.3%) | 307 (2.9%) | 163 (3.2%) |
| Vascular Access Complication | 863<br>(1.9%) | 65 (3.5%) | 102 (4.5%) | 77 (2.9%) | 78 (2.7%) | 75 (2.8%) | 67 (2.3%) | 47 (1.5%) | 60 (1.7%) | 76 (1.1%) | 128 (1.2%) | 88 (1.7%) |
| Pulmonary Vein Stenosis | 11<br>($<0.1\%$) | 0 (0%) | 3 (0.1%) | 0 (0%) | 0 (0%) | 1 ($<0.1\%$) | 1 ($<0.1\%$) | 1 ($<0.1\%$) | 1 ($<0.1\%$) | 4 ($<0.1\%$) | 0 (0%) | 0 (0%) |
| Esophageal Injury | 452<br>(1.0%) | 53 (2.9%) | 31 (1.4%) | 69 (2.6%) | 34 (1.2%) | 37 (1.4%) | 39 (1.4%) | 37 (1.2%) | 51 (1.5%) | 51 (0.7%) | 37 (0.4%) | 13 (0.3%) |
| Phrenic Nerve Injury | 16<br>($<0.1\%$) | 0 (0%) | 1 ($<0.1\%$) | 3 (0.1%) | 1 ($<0.1\%$) | 0 (0%) | 1 ($<0.1\%$) | 2 ($<0.1\%$) | 0 (0%) | 2 ($<0.1\%$) | 4 ($<0.1\%$) | 2 ($<0.1\%$) |
| Procedure-Related Arrhythmia | 1,541<br>(3.5%) | 16 (0.9%) | 28 (1.2%) | 32 (1.2%) | 37 (1.3%) | 34 (1.3%) | 108 (3.8%) | 131 (4.2%) | 136 (3.9%) | 241 (3.4%) | 462 (4.4%) | 316 (6.1%) |
| Coronary Artery Injury / Spasm | 377 (0.8%) | 19 (1.0%) | 18 (0.8%) | 16 (0.6%) | 17 (0.6%) | 14 (0.5%) | 17 (0.6%) | 24 (0.8%) | 31 (0.9%) | 46 (0.6%) | 124 (1.2%) | 51 (1.0%) |
| Hemolysis / Acute Kidney Injury | 71 (0.2%) | 0 (0%) | 0 (0%) | 0 (0%) | 0 (0%) | 0 (0%) | 0 (0%) | 0 (0%) | 0 (0%) | 11 (0.2%) | 34 (0.3%) | 26 (0.5%) |
| Respiratory / Pulmonary Complication | 367 (0.8%) | 18 (1.0%) | 34 (1.5%) | 22 (0.8%) | 20 (0.7%) | 8 (0.3%) | 22 (0.8%) | 22 (0.7%) | 22 (0.6%) | 66 (0.9%) | 86 (0.8%) | 47 (0.9%) |
| Procedure-Related Infection | 121 (0.3%) | 12 (0.7%) | 12 (0.5%) | 11 (0.4%) | 9 (0.3%) | 6 (0.2%) | 8 (0.3%) | 3 (<0.1%) | 7 (0.2%) | 17 (0.2%) | 21 (0.2%) | 15 (0.3%) |
| Procedure-Related Death | 1,355 (3.0%) | 89 (4.8%) | 106 (4.6%) | 104 (4.0%) | 108 (3.8%) | 81 (3.1%) | 92 (3.2%) | 98 (3.1%) | 139 (4.0%) | 191 (2.7%) | 235 (2.2%) | 112 (2.2%) |
| Unknown | 6 | 2 | 3 | 0 | 0 | 1 | 0 | 0 | 0 | 0 | 0 | 0 |
| Device / Equipment Malfunction | 0 (0%) | 0 (0%) | 0 (0%) | 0 (0%) | 0 (0%) | 0 (0%) | 0 (0%) | 0 (0%) | 0 (0%) | 0 (0%) | 0 (0%) | 0 (0%) |
| No Patient Harm | 16,637 (37%) | 0 (0%) | 0 (0%) | 0 (0%) | 0 (0%) | 122 (4.6%) | 850 (30%) | 1,218 (39%) | 1,411 (40%) | 4,070 (57%) | 6,376 (60%) | 2,590 (50%) |
| Other Complication | 3,072 (6.9%) | 168 (9.1%) | 188 (8.2%) | 155 (5.9%) | 138 (4.9%) | 132 (5.0%) | 207 (7.2%) | 193 (6.2%) | 244 (7.0%) | 412 (5.7%) | 709 (6.7%) | 526 (10%) |
| <sup>1</sup> n (%) |  |  |  |  |  |  |  |  |  |  |  |  |

Across 319 paired adjudications, the two models agreed on whether an event had occurred in 91.8% (Cohen’s *κ* = 0.83), and a physician reviewed the sample. Across the clinical families, the insufficient information outcome was returned for 44.2% of PFA adjudications and 44.1% of CA against 28.7% of RFA, which were not counted towards the event count.

### Disproportionality analysis

Against the reporting of other energy sources, PFA carried an ROR of 527 (95% CI, 33, 8,509) for hemolysis, 2.83 (2.55, 3.16) for arrhythmia, 2.83 (2.30, 3.48) for coronary events, 2.39 (2.10, 2.73) for cerebrovascular events, and 1.79 (1.44, 2.24) for respiratory events, each meeting the signal rule after false discovery correction (**Figure 2**). Esophageal injury, 0.024 (0.008, 0.075), pericardial events, 0.34 (0.32, 0.37), and death, 0.77 (0.67, 0.89), were reported less often with PFA. The EBGM, shrunk toward the null, met the EB05 threshold for hemolysis (4.99, EB05 = 4.08) and arrhythmia (2.16, EB05 = 2.02), while cerebrovascular events (1.99, EB05 = 1.83) and coronary events (2.23, EB05 = 1.96) sat just below it. When evaluating the quarterly reporting, as seen in **Figure 3**, 100% of hemolysis, 100% of coronary, and 80% of cerebrovascular events had an ROR exceeding 1, while 100% of esophageal and 90% of pericardial quarters sat below an ROR of 1. In comparison, the expected signal of increased risk from thermal ablation for phrenic nerve injury, pulmonary vein stenosis, esophageal damage, and pericardial events was seen consistently.

**Figure 2:**
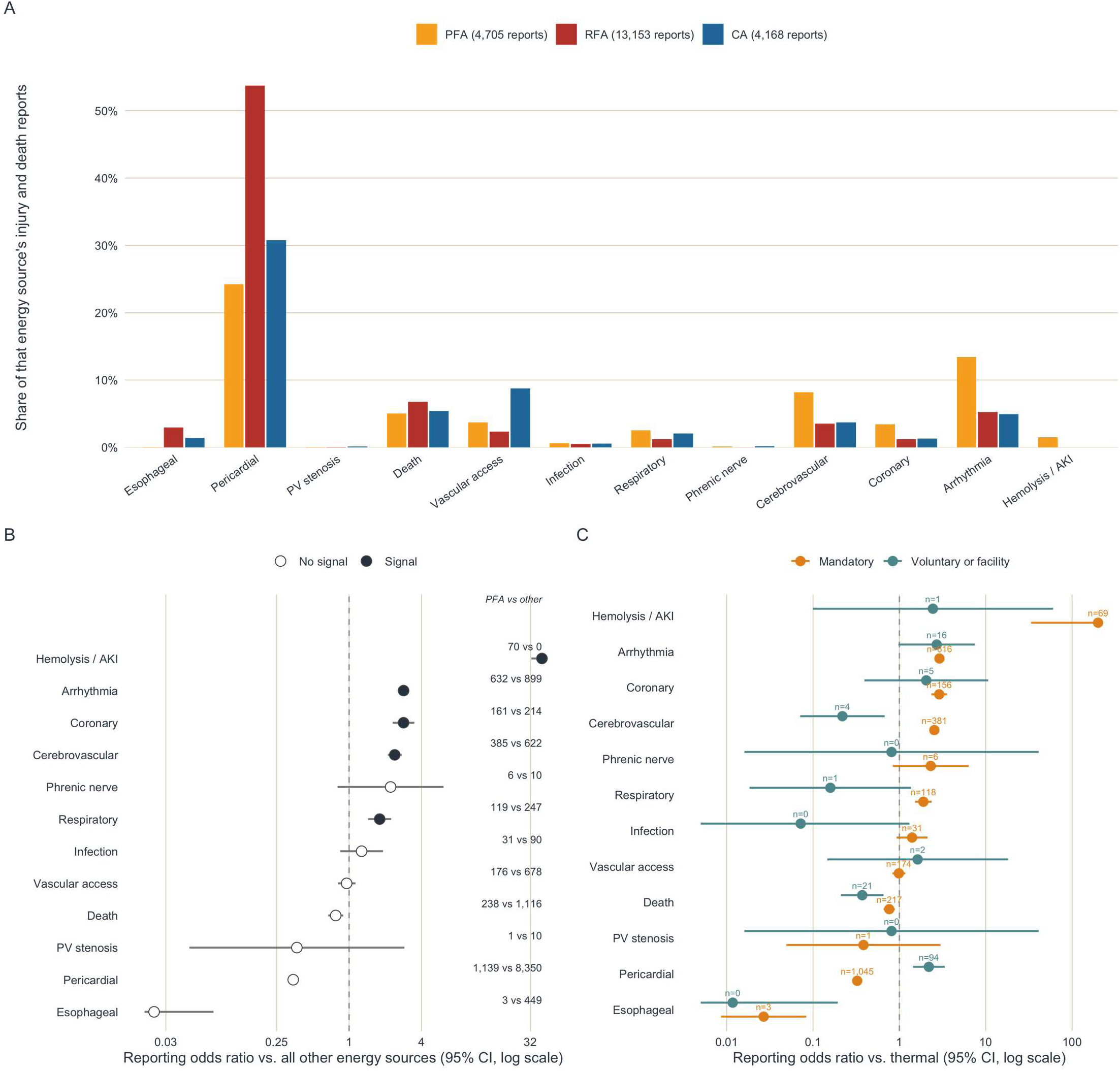
Complication reporting mix, PFA versus thermal ablation. PFA was reported in excess for hemolysis, coronary, arrhythmia and cerebrovascular events and less often for esophageal and pericardial events. (A) Confirmed complications as a share of each energy source’s own injury and death reports, a comparison of reporting mix rather than of complication rates per procedure. (B) Reporting odds ratio for each complication family, PFA versus all other energy sources, with 95% confidence intervals on a log scale; filled points met the prespecified signal rule. The hemolysis and esophageal intervals run past the axis and are pinned at the panel edge rather than dropped. (C) The same contrast by filing stream, where mandatory is by manufacturer and voluntary is by facility or physician; a family with no reports on one arm carries only the other arm’s estimate. All three panels carry the same twelve families in the same order, set by the pulsed field reporting odds ratio. AKI, acute kidney injury; CA, cryoballoon ablation; PFA, pulsed field ablation; PV, pulmonary vein; RFA, radiofrequency ablation.

**Figure 3:**
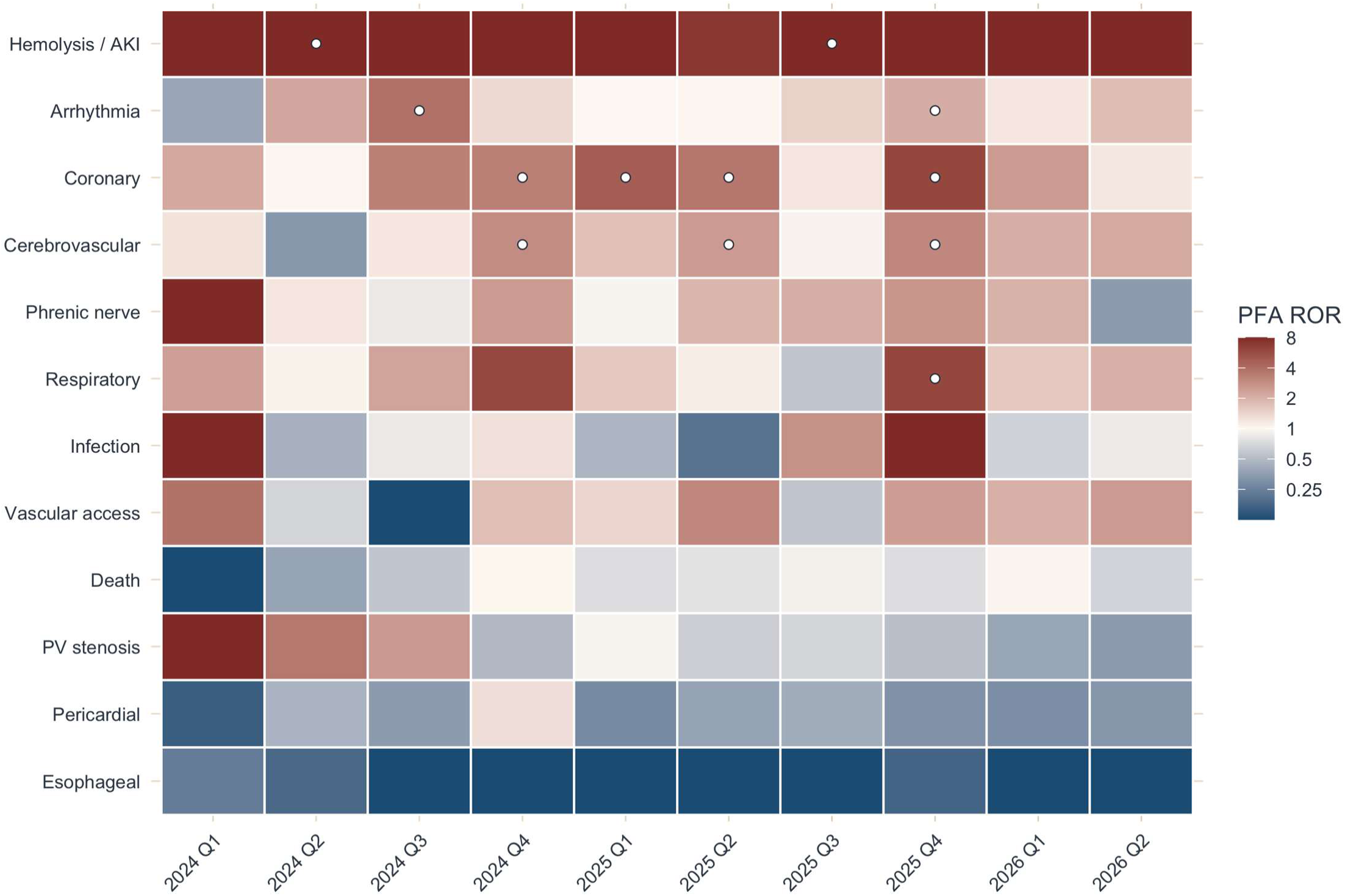
PFA reporting odds ratios by complication and quarter. Quarters start with the introduction of PFA, and dots represent false discovery rate (FDR) corrected significance. PFA, pulsed field ablation.

### Signal detection analysis

The period from 2016 through 2023, described here as the thermal era, serves as a baseline of noise for the frequency and size of series breaks, as seen in **Figure 4 (A)**. Thermal series broke 11 times across the 9 complication families available. The strongest signal was in the RFA esophageal series in January 2018 (deviance drop 39). In the PFA era, from 2024 through 2026 as seen in **Figure 4 (B)**, 7 of the 10 complication families available demonstrated a breakpoint, all between January 2024 and December 2024, with the strongest signal in cerebrovascular in December 2024 (deviance drop 40).

**Figure 4:**
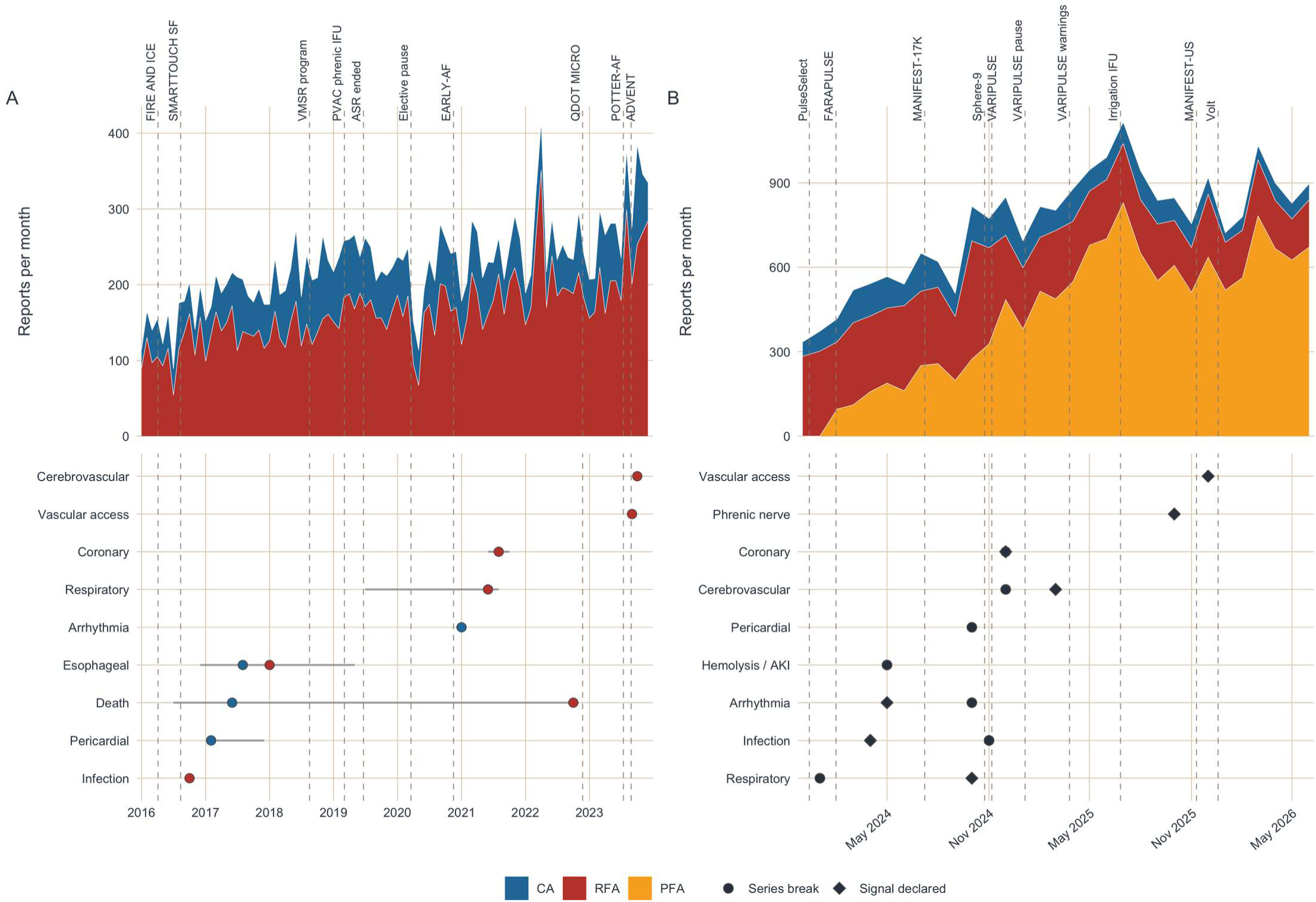
Reporting volume and signal timing in the thermal and pulsed field eras. Thermal-era series breaks set the background scale of signal noise for the pulsed field era. (A) Injury and death reports per month for radiofrequency and cryoballoon ablation from 2016 to the first pulsed field approval, and beneath it the month each complication family’s series broke under a retrospective change-point scan, with the interval around each break; a break is a change in slope, not a safety event. (B) Injury and death reports per month by energy source from the first pulsed field approval, and beneath it the month each family first broke (circle, the same scan) or was first declared (diamond, maximum sequential probability ratio test against the concurrent thermal stream). Dated approvals, safety actions and publications are marked on both panels. AKI, acute kidney injury; CA, cryoballoon ablation; IFU, instructions for use; MaxSPRT, maximized sequential probability ratio test; PFA, pulsed field ablation; RFA, radiofrequency ablation.

The prospective and retrospective signal detection methods identify different time points within the same complication family. Against concurrent thermal reporting, MaxSPRT crossed its threshold for 7 complication families, notably for cerebrovascular events in March 2025, coronary events in December 2024, and arrhythmia in May 2024. Hemolysis could not be tested, as thermal ablation carried no hemolysis reports. For cerebrovascular events, CUSUM alarmed in December 2024, the same month as detected by the break point method, while with the Shewhart method the threshold was met in May 2025. For coronary events, all detection methods signalled in December 2024. The full findings are seen in **Table S2**. Against the concurrent thermal series (**Figure 5**), the PFA reporting proportion at market entry was 5.11 (95% CI, 2.91, 8.97) times the thermal proportion for coronary events, 2.03 (1.24, 3.30) for cerebrovascular events, and 0.71 (0.51, 0.99) for pericardial events.

**Figure 5:**
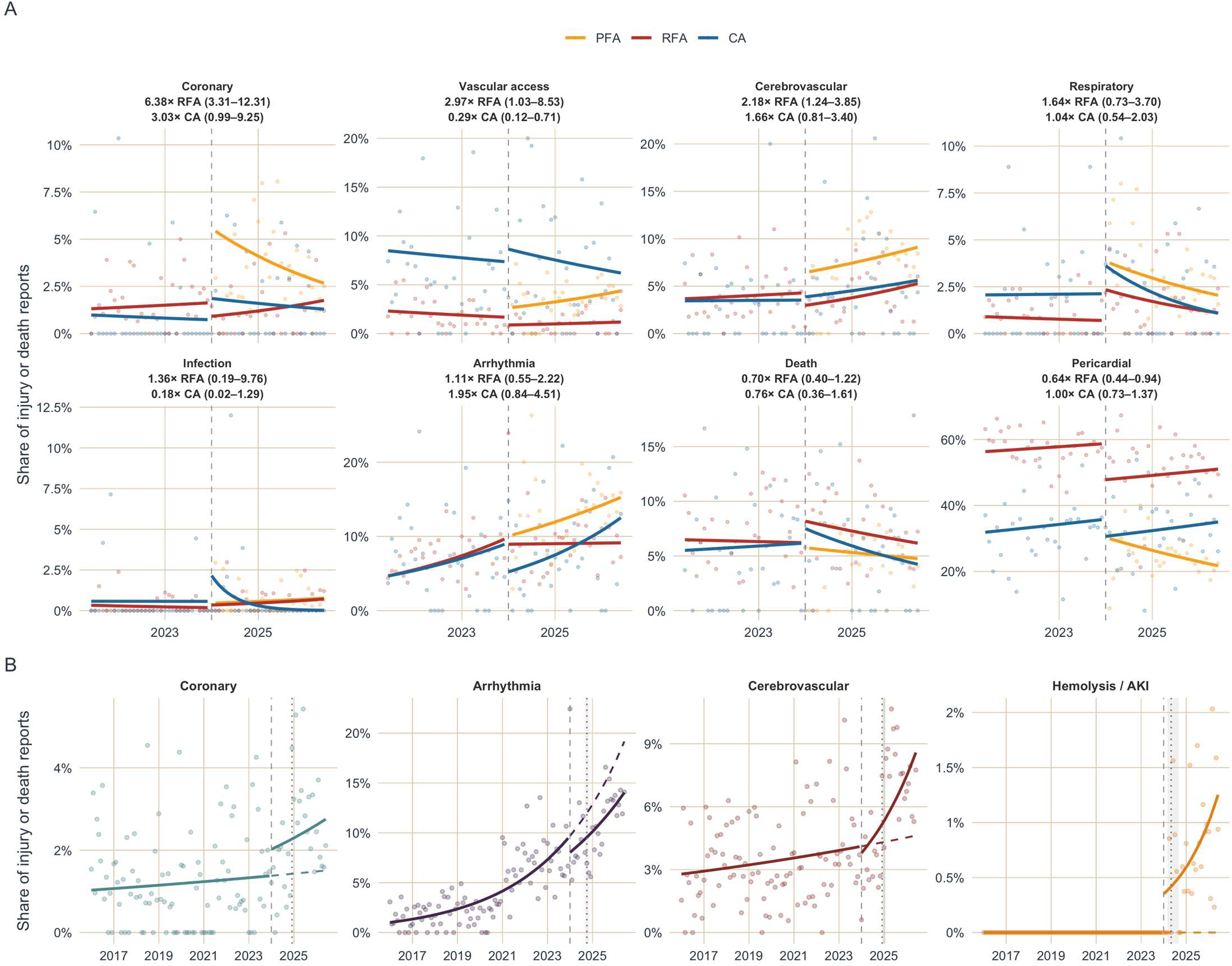
Complication reporting at PFA market entry against a concurrent thermal control. Pulsed field reporting of hemolysis, coronary, cerebrovascular and arrhythmia events increased at market entry, while pericardial and esophageal reporting decreased. (A) Monthly share of each energy source’s injury and death reports carrying each complication over the last five years, with the segmented fit for each series broken at the first pulsed field approval (dashed). Each strip carries the pulsed field level ratio against radiofrequency and against cryoballoon separately, because cryoballoon carries its own embolic profile and pooling it with radiofrequency moves the comparator. (B) Segmented quasi-Poisson fit of the pooled monthly reporting proportion for the four families of interest, with the pre-period trend carried forward as the counterfactual (dashed), the pinned break at first approval (dashed vertical) and the independently detected break with its interval (dotted, shaded). Colours are the complication palette in (B) and the modality palette in (A), and the two share no chip. CA, cryoballoon ablation; PFA, pulsed field ablation; RFA, radiofrequency ablation.

### Denominator analysis

As MAUDE provides no procedural denominator, the following results bound it under stated assumptions rather than measure it. For each complication, we determined the threshold PFA adoption rate, the minimum percentage of all AF ablations performed with PFA, at which its observed share of reports would be proportional, and therefore anticipated, under a binomial test at *α* = 0.05 (**Figure 6 (A)**). In the quarter beginning April 2026, the required share was 76.1% for cerebrovascular events, 76.1% for arrhythmia, 73.9% for vascular access, 73.5% for hemolysis, and 54.7% for coronary events, against 51.4% for pericardial and 0.0% for esophageal events, and for cerebrovascular events it had risen from 50.0% in the quarter beginning January 2025. The estimates also require the assumption that thermal reporting did not change over the same time frame. As a second bound, we computed the number of PFA procedures required to produce the observed reports at the rates published in recent large studies, if every event reached MAUDE, as demonstrated in **Figure 6 (B)**. Based on the published stroke rates, the number of PFA ablations required during 2025 would range from 38,575 by EU-PORIA to 175,618 by MANIFEST-17K, with 87,600 by EMBOL-AF, the calibration anchor.^3,4,22^ Over the same time frame, based on the published rates of coronary events, the number required would range from 3,607 by NEMESIS-PFA to 163,333 by MANIFEST-PF.^2,17^

**Figure 6:**
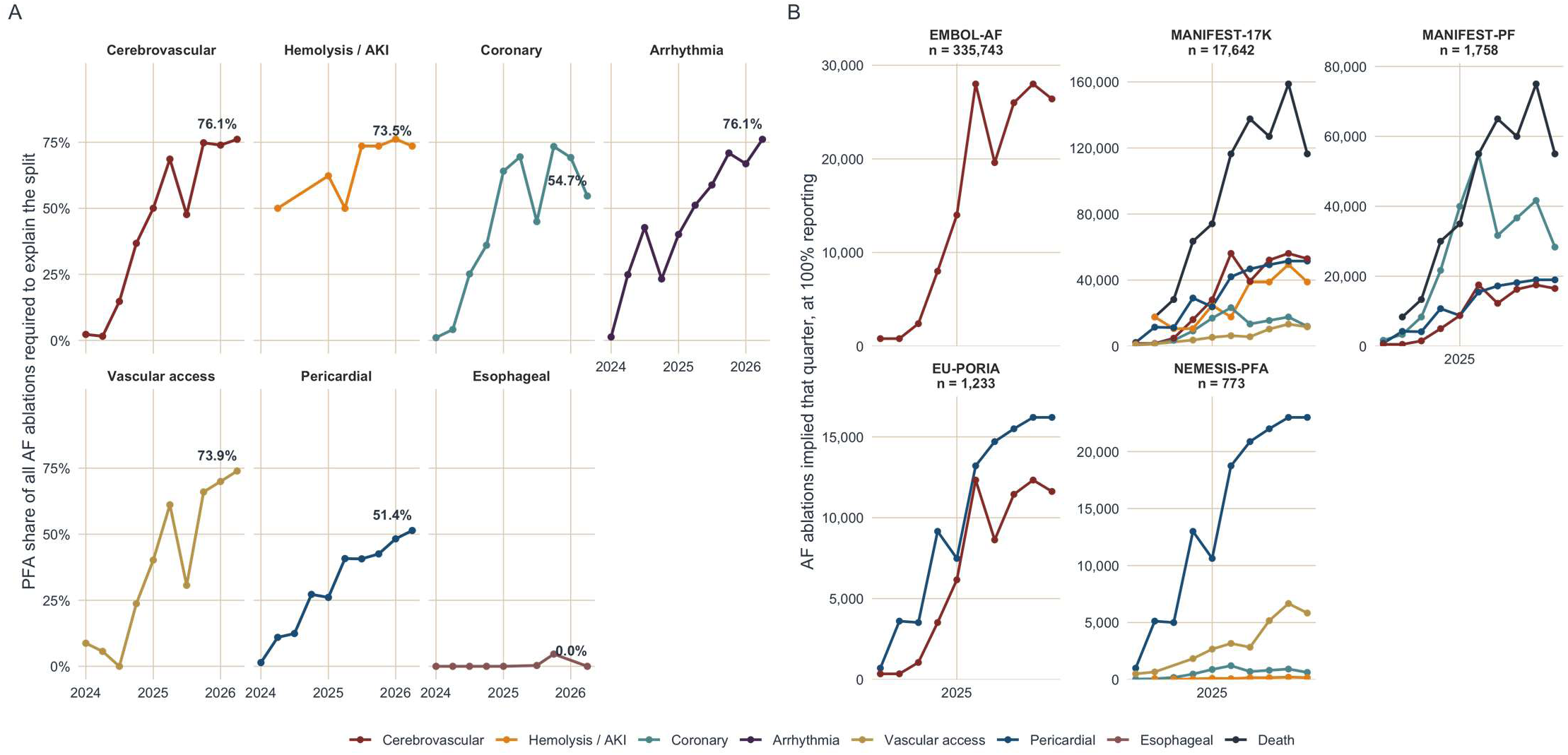
Procedure volume bounds on PFA complication reports. Procedure volume alone cannot explain the pulsed field share of cerebrovascular, hemolysis, coronary and arrhythmia reports, and under EMBOL-AF, the calibration anchor, the implied procedure volumes fall below plausible real-world volumes, indicating that MAUDE captures a minority of events. (A) The smallest PFA share of procedures in each quarter at which the observed PFA share of reports would no longer exceed expectation under a binomial test at α = 0.05; the last quarter is labelled, and quarters with fewer than five reports are omitted. (B) Pulsed field procedures implied in each quarter if every event reached MAUDE, computed as confirmed reports divided by each study’s published rate, with panels ordered by cohort size on a free y-axis. Rates from EMBOL-AF,^22^ MANIFEST-17K,^3^ MANIFEST-PF,^2^ EU-PORIA,^4^ and NEMESIS-PFA.^17^ AKI, acute kidney injury; MAUDE, Manufacturer and User Facility Device Experience; PFA, pulsed field ablation.

### Sensitivity analyses

We additionally broke down events by complication type and by platform. As shown in **Figure 7**, the cerebrovascular ROR was highest for VARIPULSE (14.11 (95% CI, 10.93, 18.21)), followed by Affera Sphere-9 (1.59 (1.12, 2.28)) and FARAPULSE (1.39 (1.12, 1.72)), while PulseSelect (1.28 (0.88, 1.87)) was not. With VARIPULSE removed, the class PRR fell from 2.01 (1.69, 2.39) to 1.36 (1.13, 1.65) and stayed above 1 (**Figure 7**). After the VARIPULSE IFU change, there was a decrease in the proportion of cerebrovascular event reports, with no change seen to other platforms (**Figure S1**). A break down by severity tiers is shown in **Figure 7**. The pooled cerebrovascular series broke in December 2024 and MaxSPRT crossed in March 2025, 13 months before the *AVANT GUARD* neurologic events were published in April 2026,^19^ and across that break the FARAPULSE cerebrovascular share rose from 2.9% to 6.1% of its injury or death reports.

**Figure 7:**
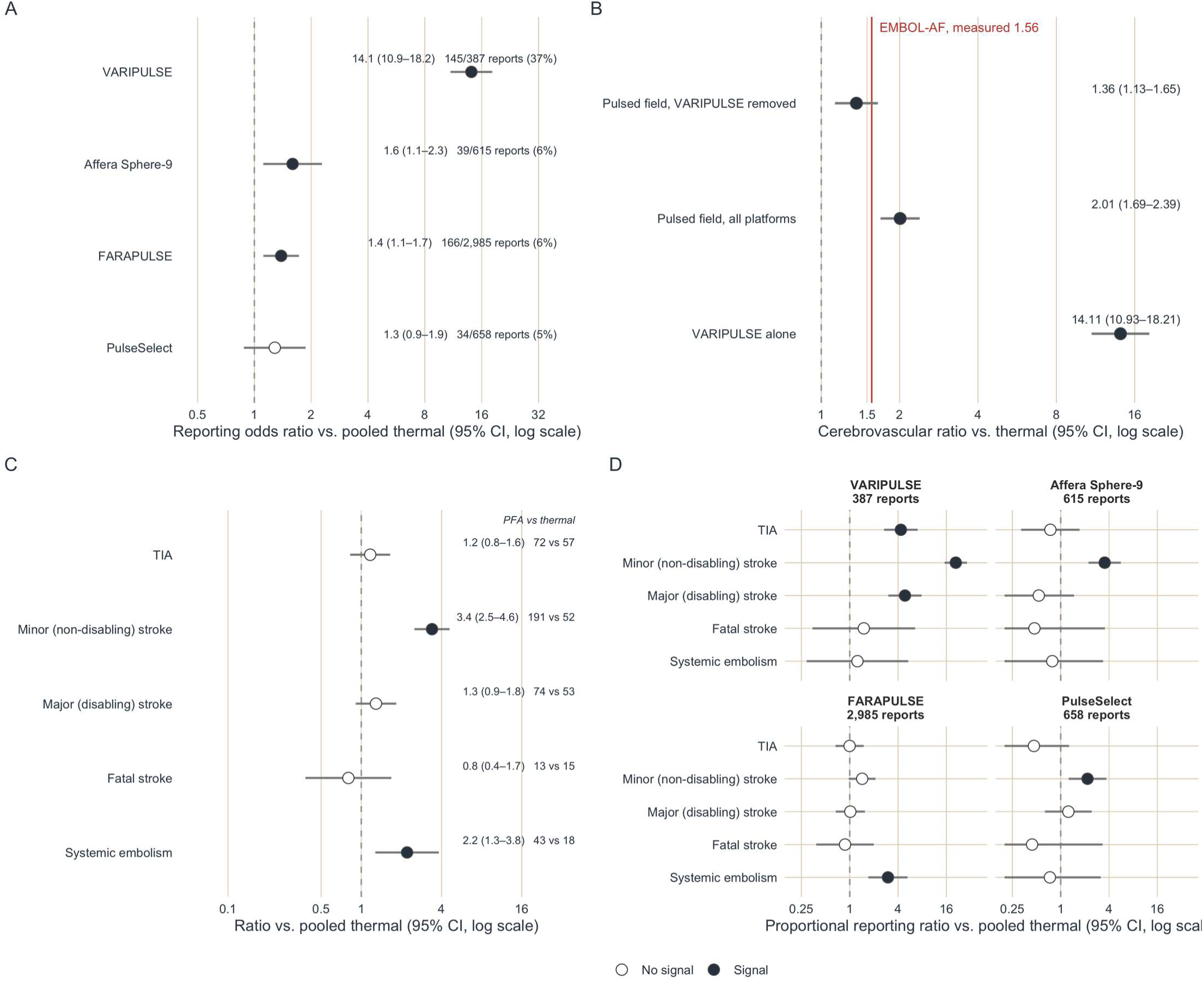
Cerebrovascular reporting by PFA platform and severity tier. The excess is largest for VARIPULSE but is also seen with other PFA platforms. With VARIPULSE removed the class estimate falls but stays above 1, bracketing the ratio EMBOL-AF measured, with the excess seen in the minor-stroke and systemic-embolism tiers. (A) Cerebrovascular reporting odds ratio for each pulsed field platform against pooled thermal ablation, with the platform’s confirmed events over its injury and death reports. (B) The proportional reporting ratio for the class with every platform and with VARIPULSE removed, and the reporting odds ratio for VARIPULSE alone, beside the pulsed field to thermal incidence ratio measured in the EMBOL-AF registry (vertical line). (C) Pulsed field against pooled thermal within each adjudicated cerebrovascular severity tier; filled points met the signal rule. (D) The same severity contrast within each platform, harder to interpret due to decreased numerators, and classification biased by manufacturer reporting style. CI, confidence interval; PFA, pulsed field ablation; TIA, transient ischemic attack.

Re-estimated on reports received from January through June 2026 alone, against estimates from reports received through December 2025, the cerebrovascular, arrhythmia and hemolysis signals replicated, and coronary events kept their direction at 1.74 (95% CI, 0.84, 3.59) without meeting the signal rule (**Figure S2**).

## DISCUSSION

In this analysis of the 44,623 MAUDE reports across three energy modalities from 2016 through 2026, we found that PFA carried a reporting excess for hemolysis, cerebrovascular, coronary, and arrhythmic events against thermal ablation, and a deficit for esophageal and pericardial events. We found that the excess complication risk also mirrored increases in event rates in active trials and registries, preceding their publications by months to years, and were specific to platforms and complication types. Hemolysis and related kidney injury remain the most disproportionately reported event, across PFA platforms, and continues into 2026 without significant change in reporting frequency. Cerebrovascular events were reported in excess on every platform except PulseSelect, and the class PRR of 2.01 (95% CI, 1.69, 2.39) remained elevated at 1.36 (1.13, 1.65) even after removal of VARIPULSE, concordant in size and direction with the ratio of 1.56 measured in *EMBOL-AF*.^22^ Every complication family that later carried a signal demonstrated a break point within the first year of approval, and sequential testing showed that platform-specific and severity-specific signal excess were corroborated by hold-out data from 2026. Together, we demonstrated the utility of a prospective signal detection method over that of any one disproportionality analysis, supported by an automated adjudication system, can reveal signal breaks by modality and platform and allow for validation against different historical eras and time points.

With tens of thousands of reports each year, MAUDE can no longer be adjudicated by hand. While some have managed to manually adjudicate small subsets of MAUDE event narratives, most studies are based off of term matching and classification.^20,24,39,40^ LLM-guided adjudication carries a risk of systematic error, but it offers a powerful means of classifying large datasets with greater speed and consistency than manual review. There are inconsistencies inherent to the MAUDE database, including differences between manufacturers, voluntary and mandatory reporting styles, and the details of the clinical event itself. For example, although tamponade was reported less often with PFA, our analysis detected a signal for an increased reporting rate of pericarditis, two events that might otherwise have been conflated under a single “pericardial injury” term (**Figure S3**).^3,41,42^ Within arrhythmia events, additional need for cardioversion, drug therapy, or repeat ablation were noted in the PFA group, while in the RFA group, permanent device implantation was more common (**Figure S3**). On the other hand, coronary events were confined to only transient spasm, with no excess in sustained spasm, infarction, or stenosis, consistent with the described mechanism.^14–16^ We were also able to classify report types by platform in addition to event severity. For example, the cerebrovascular event signal of PFA was previously attributed to VARIPULSE alone when compared against other PFA platforms.^20,40^ In our review, although VARIPULSE had an increased ROR of 14.11 (95% CI, 10.93, 18.21) for cerebrovascular events, the other platforms, with the exception of PulseSelect, still demonstrated increased risk over thermal ablation (**Figure 7**). The revised VARIPULSE protocol reports fewer complications,^43^ which we also note after the change in the IFU in June 2025 [**Figure S1**], however compared to the thermal counterparts, the reporting risk remained elevated, although confounded by notoriety bias. Similarly, the *AVANT GUARD* trial noted an early increase in stroke rates with FARAPULSE, which was identifiable in the MAUDE database significantly prior to publication of the trial results - that type of safety signal could potentially be used prospectively to modify practice in response, such as increasing anticoagulation levels as part of the procedural protocol. These signals can be slower to surface in industry-sponsored Investigational Device Exemption trials, as these are usually designed with composite safety endpoints rather than specific individual safety endpoints.^8^

Several limitations need to be considered for our study. One important limitation with the interpretation of MAUDE data is the lack of a procedural denominator, which we attempted to resolve by providing theoretical rates and denominators. The rationale behind this was to assess whether the introduction of PFA was associated with increased adverse event profiles in the real-world experience, expressing reporting excesses as the PFA usage share or procedural volume it would imply, rather than to quantify specific risk rates. By inverting the binomial disproportionality test, we were able to provide an evaluation of the minimum share of total ablations using PFA that would be required to statistically explain the volume of MAUDE reports attributed to PFA (**Figure 6**). This circumvents the need for estimates of the distribution of procedures across energy sources, which are not reliably known. For example, by the quarter beginning April 2026, at least 76.1% of ablations would have had to be with PFA to explain the excess share of cerebrovascular events over that of thermal. Registry data place PFA adoption at roughly 60% in 2025 among centers reporting to the National Cardiovascular Data Registry (**NCDR**),^44^ and those centers are among the earliest adopters, so national adoption is unlikely to have exceeded that. Procedure mix alone therefore does not account for the excess.

Another major issue is underreporting of events, both in MAUDE and in the NCDR database. We believe that underreporting bias likely strengthens our findings, as the events captured in the MAUDE database are likely an underrepresentation of the real world experience. To bound the procedure volume these reports imply, we applied the reported complication rates in large studies to the number of PFA event reports, assuming 100% event reporting, to estimate the PFA procedure volume that would have to have been performed for that event rate to hold true. Where the implied volume exceeds the number of procedures plausibly performed, the published literature may be underestimating complication rates, and where it falls short, the gap is more likely evidence of underreporting to MAUDE. This approach allows us to bound the denominator for emerging technologies before procedure-level data are available.

The record is also incomplete and noisy, and different statistical approaches flagged the same complication family at different times or not at all (**Table S2**), leaving open which alarm to act on prospectively. Additionally, manual adjudication of a record this large would be extremely time-intensive, so we used an LLM, whose accuracy across the full record will be imperfect despite a second-model check and physician review of a sample. These limitations guide the next steps, from improved adjudication methods to prospective implementation of surveillance in MAUDE.

## Conclusion

In this MAUDE analysis, PFA showed disproportionate reporting of hemolysis, cerebrovascular, coronary, and arrhythmic events, and the cerebrovascular, arrhythmia and hemolysis excesses replicated in reports received from January through June 2026. Large language model adjudication of report narratives made prospective signal detection feasible at scale, surfacing the cerebrovascular signal months before *AVANT GUARD* published it and suggesting a scalable tool for early detection of procedural risk as new ablation technologies enter practice. We believe that combining this data with clinical trials and registry data will allow us to minimize the exposure of risk to patients.

## Data Availability

All data and code used for analysis is available to the public through open-source software and public datasets.

https://cran.r-project.org/package=card

https://www.accessdata.fda.gov/scripts/cdrh/cfdocs/cfmaude/search.cfm

## NON-STANDARD ABBREVIATIONS AND ACRONYMS

AF: Atrial fibrillation
CA: Cryoballoon ablation
CUSUM: Cumulative sum
EB05: Empirical Bayes 5th percentile
EBGM: Empirical Bayes geometric mean
FDA: Food and Drug Administration
IFU: Instructions for use
LLM: Large language model
MAUDE: Manufacturer and User Facility Device Experience
MaxSPRT: Maximized sequential probability ratio test
NCDR: National Cardiovascular Data Registry
PFA: Pulsed field ablation
PRR: Proportional reporting ratio
RFA: Radiofrequency ablation
ROR: Reporting odds ratio

## SUPPLEMENTAL MATERIAL

- Supplemental Methods: data source, device problems, event adjudication, disproportionality, and interrupted time series and signal detection
- Supplemental Results: report characteristics, complication mix by energy source, device problems, severity and platform-specific analysis, and calibration and sensitivity analyses
- **Table S1**: Complication families and severity tiers
- **Table S2**: Earliest month each signal detection method fired
- **Figure S1**: VARIPULSE cerebrovascular reporting before and after the irrigation instructions for use were revised
- **Figure S2**: Hold-out re-estimation on reports from January to June 2026
- **Figure S3**: PFA reporting by severity tier within complication families
- **Figure S4**: Device problems in malfunction reports
- Statistical Software: R version, platform, and package versions

## Supplemental Methods

### Data source

We searched five product codes that encompassed the catheters used for AF ablation: QZI (percutaneous cardiac ablation catheter for atrial fibrillation with irreversible electroporation), OAE (percutaneous cardiac ablation catheter for atrial fibrillation), LPB (cardiac ablation percutaneous catheter), OAD (percutaneous cardiac ablation catheter for atrial flutter), and DRA (steerable catheter). All reports were carried as a sensitivity denominator to assess overall reporting rates.

### Device problems

We took the International Medical Device Regulators Forum (**IMDRF**) Annex A device problem terms coded on each malfunction report, grouped them by their Annex A family term, and compared each energy source’s problem mentions before and after PFA approval.

### Event adjudication

We paired each family with the IMDRF patient problem terms by matching the full term list through manual physician review. The LLM was given the same prompt and instructions, and defaulted to absent unless the narrative supported the tier, with an insufficient information outcome retained as an alternative. The adversarial sample was stratified by complication type, and GPT-5.6 Luna was given the same definitions and tiers. A physician reviewed the sample for the event labels and complication tiers.

### Disproportionality

The information component is a Bayesian log_2_ of observed to expected reports, and the EBGM shrinks the relative reporting ratio towards the null when a cell carries little data.^30,31^

### Interrupted time series and signal detection

Quasi-Poisson models were fitted on monthly counts to maintain point estimates while accounting for highly dispersed data. Diagnostics were performed on each interrupted time series fit, including lag autocorrelation and a Ljung-Box test of residuals, F test on seasonality, and negative binomial refit to assess variance assumptions. Complications with too few events for signal detection were reported as not estimable.

## Supplemental Results

### Report characteristics

The malfunction reports were 65.0% of PFA reports against 42.0% of RFA and 50.9% of CA. The median time from event to FDA report was 20 to 28 days, and 47.1% of PFA reports originated in the United States against 39.7% of RFA and 31.8% of CA. Manufacturers, as mandatory reporters, filed 96.4% to 98.9% of each modality’s reports, and voluntary reports held 243 PFA reports against 197 thermal reports (**Figure 2 (C)**).

### Complication mix by energy source

Within each modality, as seen in **Figure 2 (A)**, cerebrovascular events were 8.2% of PFA reports against 3.6% of RFA and 3.7% of CA, arrhythmia 13.4% against 5.3% and 5.0%, and coronary events 3.4% against 1.2% and 1.3%, with hemolysis reported by PFA alone. Pericardial events were 24% of PFA reports against 54% of RFA and 31% of CA, and esophageal injury 0.06% against 3.0% and 1.4%.

### Device problems

In the malfunction reports, PFA had 7,630 IMDRF device problem reports compared to the 4,497 for RFA (**Figure S4**). The most common device related problem families were material integrity issues (25.7%), followed by device positioning/separating problems (19.5%). The most common specific problems were entrapment of the device itself or material deformation.

### Severity and platform-specific analysis

Within families (**Figure S3**), the cerebrovascular excess sat in minor non-disabling stroke, 3.39 (95% CI, 2.50, 4.60) on 191 PFA against 52 thermal events, and systemic embolism, 2.21 (1.27, 3.82) on 43 against 18, with transient ischemic attack (**TIA**) 1.17 (0.83, 1.65), major disabling stroke 1.29 (0.91, 1.83), and fatal stroke 0.80 (0.38, 1.68) null; a tier could not be assigned in 501 PFA and 306 thermal reports, 1.51 (1.32, 1.73), as imaging and follow-up were often absent from the narrative. The coronary excess was confined to transient spasm, 10.47 (5.30, 20.66) on 102 against 9, with sustained spasm 2.77 (0.89, 8.58), myocardial infarction 0.99 (0.65, 1.50), and coronary stenosis 0.35 (0.09, 1.30) null. Every mechanical pericardial tier ran below 1, tamponade treated by pericardiocentesis at 0.48 (0.44, 0.51) and by surgery at 0.56 (0.48, 0.65), while pericarditis without effusion ran the other way, 2.22 (1.36, 3.65) on 53 against 22. Within arrhythmia, self-terminating events 1.91 (1.41, 2.57), events requiring cardioversion or drugs 2.78 (2.25, 3.43), and repeat ablation 1.87 (1.32, 2.64) were reported more often with PFA, and a permanent pacemaker or defibrillator less often, 0.49 (0.39, 0.63) on 94 against 176. All 70 confirmed hemolysis events were PFA, 15 laboratory only, 31 with acute kidney injury without dialysis, and 24 requiring dialysis.

### Calibration and sensitivity analyses

The reference set of expected associations recovered 6 of 6 expected thermal excesses, 1 of 3 expected PFA deficits, and 3 of 6 expected nulls. The 3 null pairs in which no other modality was expected to signal had a geometric mean ROR of 1.03 (SD of the log ROR, 0.23), against 0.68 (1.04) across all 6 expected nulls, too few pairs to fit a calibration distribution, so the estimates were reported uncalibrated. The information component lower bound exceeded 0 for cerebrovascular, arrhythmia, coronary, hemolysis and respiratory events, including cerebrovascular events at 0.84 (0.69, 0.98). With all reports as the denominator rather than injury or death reports alone, the PFA ROR was lower by a factor of 1.35 to 1.70 across families, signal status changed only for respiratory events, and MaxSPRT crossed for cerebrovascular events in February 2026 rather than March 2025. Across the pooled interrupted time series fits, Ljung-Box tests indicated residual autocorrelation for arrhythmia, coronary and hemolysis events, the F test indicated seasonality for infection events, and a negative binomial refit moved no level estimate by more than 12%.

## Supplemental Tables

**Table S1:** Complication families and severity tiers as given to the adjudication model. Definitions are reproduced from the schema the model was prompted with, so the table cannot drift from what was actually adjudicated. insufficient_info is an adjudication outcome rather than a severity and is never ordered with the tiers above it.

| Severity tier | Definition |
| --- | --- |
| Pericardial Effusion / Cardiac Tamponade / Pericarditis. Pericardial complication during or after AF catheter ablation: effusion (blood or serous), frank tamponade, or post-ablation pericarditis (with or without effusion). Mechanisms: atrial wall or transseptal-needle perforation, thermal injury with delayed rupture, over-anticoagulation, inflammatory pericarditis. Cues: hypotension, pulsus paradoxus, pleuritic chest pain, friction rub, effusion on echo, pericardiocentesis, surgical drainage, NSAIDs/colchicine. Includes acute intraprocedural and delayed (hours to days) presentations. |  |
| Pericarditis, no effusion | Pericarditis (pleuritic chest pain, possible friction rub) without significant effusion. Managed with NSAIDs, colchicine, or observation. |
| Trivial effusion | Small or trace effusion at end of case or on post-procedure imaging. No hemodynamic compromise, no drainage. Observation with or without anti-inflammatories. |
| Moderate, no intervention | Moderate effusion with symptoms (chest pain, dyspnea, tachycardia) but no drainage. May include holding or reversing anticoagulation and medical therapy. |
| Tamponade, pericardiocentesis | Hemodynamically significant effusion or tamponade requiring percutaneous pericardiocentesis. Hypotension, pulsus paradoxus, or chamber collapse prompting emergent drainage. |
| Tamponade, surgical | Tamponade or perforation requiring surgery: pericardial window, thoracotomy, sternotomy, or atrial wall repair. |
| Reported, severity unclear | Pericardial effusion, tamponade, or pericarditis mentioned but detail insufficient to determine intervention or management. |
| Cerebrovascular Event. Ischemic stroke, hemorrhagic stroke, transient ischemic attack (TIA), or systemic thromboembolism during or after AF ablation. Mechanisms: thrombus on catheters or sheaths, air embolism (transseptal puncture, catheter exchange), char embolism, dislodged pre-existing left atrial thrombus. Cues: focal deficit (weakness, aphasia, visual change), altered mental status, infarct or hemorrhage on CT/MRI, systemic embolism. Includes clinically apparent events and silent (asymptomatic) cerebral lesions on post-procedure DWI-MRI. |  |
| TIA | Transient focal deficit fully resolving within 24 hours with no infarct on imaging. Return to neurologic baseline. |
| Minor (non-disabling) stroke | Non-disabling stroke: mild residual deficit at discharge or follow-up (modified Rankin scale <2, or NIHSS <=3), or labeled 'minor stroke.' Also silent cerebral lesions on post-procedure MRI, if reported. |
| Major (disabling) stroke | Disabling stroke: persistent deficit with lasting functional disability (modified Rankin scale >=2 attributable to the event), ICU-level care, or neurosurgical intervention. Includes large-territory infarction and symptomatic intracranial hemorrhage. |
| Fatal stroke | Cerebrovascular event that directly caused or substantially contributed to death. |
| Systemic embolism | Thromboembolism to a non-cerebral territory (mesenteric, renal, or peripheral arterial) in the peri-procedural period. |
| Reported, severity unclear | Cerebrovascular or embolic event described but deficit severity, symptom duration, or outcome unclear. |
| Vascular Access Complication. Complication of percutaneous vascular access, usually at the femoral venous or arterial puncture site. Cues: groin hematoma, swelling, access-site pain, pseudoaneurysm, arteriovenous (AV) fistula, retroperitoneal hemorrhage, or bleeding requiring transfusion. Also access-related deep vein thrombosis or pulmonary embolism and catheter/sheath vascular injury. EXCLUDES bleeding at non-access sites. |  |
| Minor hematoma | Groin hematoma or minor access-site bleeding managed with compression, observation, or bed rest only. No transfusion or procedural repair. |
| Major hematoma transfusion | Access-site hematoma or bleeding requiring transfusion but not surgical or interventional repair. Includes hemoglobin drop of 2 g/dL or more. |
| Pseudoaneurysm | Femoral pseudoaneurysm requiring intervention: ultrasound-guided thrombin injection, compression, or surgical repair. |
| Av fistula | Access-site arteriovenous fistula. Managed by observation, compression, or surgical repair per size and symptoms. |
| Retroperitoneal | Retroperitoneal hemorrhage from femoral vessel injury. Flank or abdominal pain, hemodynamic instability, hemoglobin drop. May require transfusion, embolization, or surgery. |
| Vte | Access- or catheter-related deep vein thrombosis or pulmonary embolism. |
| Reported, severity unclear | Vascular access complication described but type or severity unclear. |
| Pulmonary Vein Stenosis. Narrowing of one or more pulmonary veins from ablation within or at the PV ostium. Incidence fell substantially with the shift from ostial to antral isolation. Seen with radiofrequency and cryoballoon; rare with pulsed field ablation (tissue selectivity). Cues: dyspnea, hemoptysis, recurrent pulmonary infection, PV narrowing on CT/MRI. Onset often weeks to months after ablation. |  |
| Asymptomatic | PV narrowing on follow-up imaging (CT, MRI, or TEE), patient asymptomatic. Includes mild (<50% luminal reduction) and moderate (50-70%) stenosis without symptoms. |
| Symptomatic medical | Symptomatic PV stenosis (dyspnea, hemoptysis, recurrent pneumonia, reduced exercise tolerance) managed medically or by observation without intervention. |
| Intervention required | PV stenosis requiring balloon angioplasty or stent. Usually severe (>70% luminal reduction) with limiting symptoms. |
| Complete occlusion | Complete occlusion of one or more pulmonary veins on imaging. May be asymptomatic with adequate collaterals; most severe anatomic finding. |
| Reported, severity unclear | PV stenosis or related symptoms mentioned but imaging, severity, or management insufficient to grade. |
| Esophageal Injury. Esophageal injury from energy delivery on the posterior left atrial wall (adjacent to the anterior esophagus). Spectrum: superficial mucosal injury to the catastrophic, often fatal atrioesophageal fistula (AEF). All energy modalities; lower thermal risk with PFA (tissue selectivity). Cues: chest or back pain, dysphagia, odynophagia, fever, endoscopic erythema or ulceration, mediastinal air on CT, neurologic symptoms from air embolism via fistula. Also periesophageal vagal injury causing gastroparesis: nausea, vomiting, early satiety, bloating. |  |
| Mucosal injury | Superficial injury limited to mucosal erythema, erosion, or shallow ulcer on endoscopy. Managed with proton pump inhibitors and diet. No perforation or fistula. |
| Deep ulceration | Deep ulceration beyond the mucosa without fistula. Extended medical management (sucralfate, IV PPI) and surveillance imaging. |
| Atrioesophageal fistula | AEF confirmed on imaging (mediastinal or left atrial air on CT) or at surgery. Life-threatening; typically 2-5 weeks post-ablation with fever, neurologic symptoms from air or septic embolism, hematemesis, or sepsis. |
| Gastroparesis | Symptomatic gastroparesis (nausea, vomiting, early satiety, bloating, delayed gastric emptying) from periesophageal vagal injury during posterior wall ablation. Self-limited to debilitating. |
| Reported, severity unclear | Esophageal symptoms, findings, or concern described but injury depth or outcome unclear. |
| Phrenic Nerve Injury. Right or left phrenic nerve injury during ablation causing diaphragmatic paresis or paralysis. Right nerve most affected (proximity to right superior PV and superior vena cava). Most common with cryoballoon of the right superior PV; also radiofrequency, rarely PFA. Cues: loss of diaphragmatic excursion during the procedure (fluoroscopic or pacing-monitored), post-procedure dyspnea, elevated hemidiaphragm on chest X-ray, reduced inspiratory effort. |  |
| Intraprocedural only | Phrenic capture lost or reduced diaphragmatic excursion noted intraprocedurally, prompting ablation cessation. Function recovered before end of case or by discharge. |
| Transient | Palsy persisting beyond the procedure but recovering fully within 12 months. Includes elevated hemidiaphragm at discharge with later recovery. |
| Persistent | Palsy still present at 12 months or later, or described as permanent. May have chronic exertional dyspnea and persistent hemidiaphragm elevation. |
| Reported, severity unclear | Phrenic injury or elevated hemidiaphragm described but follow-up insufficient to determine recovery. |
| Procedure-Related Arrhythmia. New arrhythmia caused by the ablation itself, distinct from recurrence of the original AF. Includes iatrogenic left atrial macro-reentrant tachycardia or atypical flutter from lesion-set gaps, new AV block from septal ablation or conduction-system trauma, inappropriate sinus tachycardia from autonomic modulation, and new ventricular or organized atrial proarrhythmia. EXCLUDES recurrent pre-existing AF and expected blanking-period reconnection arrhythmias. |  |
| Self-terminating | Iatrogenic arrhythmia terminating spontaneously or with brief pacing maneuvers, no recurrence, no added intervention. |
| Cardioversion or drugs | Requires electrical cardioversion, antiarrhythmic, or rate-control drugs. Includes persistent atrial tachycardia or flutter managed medically without repeat ablation. |
| Repeat ablation | Requires a repeat ablation (e.g., mapping and ablation of a gap-related macro-reentrant tachycardia or flutter circuit). |
| Pacemaker or ICD | Requires a permanent pacemaker or ICD. Includes complete AV block from septal ablation or catheter trauma needing permanent pacing. |
| Reported, severity unclear | New ablation-related arrhythmia described but type, management, or outcome unclear. |
| Coronary Artery Injury / Spasm. Coronary artery spasm, occlusion, or injury from ablation energy near the coronaries. Most reported with pulsed field ablation (PFA), whose high-voltage field can stimulate vascular smooth muscle: focal left circumflex spasm during mitral isthmus ablation and right coronary spasm during cavotricuspid isthmus ablation are the recognized patterns. Delayed diffuse spasm (possibly hemolysis-related nitric oxide depletion) also described with PFA. Cues: ST-segment changes, chest pain, hemodynamic instability, angiography findings, intraprocedural nitroglycerin. Rare with radiofrequency, mainly epicardial ablation. |  |
| Transient spasm | Spasm resolving promptly with cessation of energy and/or intracoronary or IV nitroglycerin. No myocardial injury (normal troponin). No lasting ischemia. |
| Sustained spasm | Spasm requiring prolonged vasodilators, occurring remote/delayed from energy delivery, or recurring after resolution. May include troponin rise not meeting MI criteria. |
| Myocardial infarction | Acute MI from occlusion or sustained spasm. Includes STEMI, NSTEMI with significant troponin rise plus ischemic symptoms or ECG changes, and occlusion requiring emergent PCI. |
| Coronary stenosis | Coronary narrowing or remodeling on follow-up angiography or OCT attributed to ablation injury. Recently described with PFA near coronary vessels. |
| Reported, severity unclear | Coronary symptoms, ST changes, or concern described but mechanism, severity, or outcome unclear. |
| Hemolysis / Acute Kidney Injury. Intravascular hemolysis from electroporation of red blood cells, predominantly with pulsed field ablation (PFA). High-voltage pulsed fields rupture erythrocytes (transmembrane pore formation, osmotic swelling). Cues: elevated plasma free hemoglobin, elevated LDH, hemoglobinuria (dark urine), and in severe cases acute kidney injury (AKI) from heme-mediated proximal tubular damage. Free hemoglobin also scavenges nitric oxide (may contribute to urinary retention, coronary spasm, hypertension). Risk rises with number of PFA applications and catheter-tissue contact. Severe AKI requiring dialysis is rare (~0.03%, MANIFEST-17K). |  |
| Laboratory only | Lab evidence of hemolysis (elevated free hemoglobin, LDH, hemoglobinuria) without significant renal dysfunction or other end-organ effect. Self-resolving within 24-48 hours. |
| AKI, no dialysis | Hemolysis-induced AKI (significant serum creatinine rise) managed conservatively with IV hydration, no renal replacement. |
| AKI requiring dialysis | Severe hemolysis-induced AKI requiring temporary or sustained renal replacement therapy (hemodialysis). |
| Reported, severity unclear | Hemolysis, dark urine, elevated LDH, or renal dysfunction mentioned but severity unclear. |
| Respiratory / Pulmonary Complication. Pulmonary or thoracic complication NOT from pulmonary vein stenosis (see pv_stenosis) or phrenic nerve injury (see phrenic). Includes pneumothorax (subclavian, internal jugular, or epicardial access), hemothorax, pulmonary hemorrhage or hemoptysis (reported with PFA), bronchial injury, pulmonary edema, and pulmonary infection. Cues: dyspnea, chest pain, reduced breath sounds, chest tube placement, chest imaging findings. |  |
| Mild | Minor, managed conservatively: small pneumothorax resolving without chest tube, transient hemoptysis, or mild pulmonary edema treated with diuretics. |
| Moderate intervention | Requires procedure: chest tube for pneumothorax or hemothorax, or significant pulmonary hemorrhage requiring bronchoscopy. |
| Severe | Requires ICU care, mechanical ventilation, surgery, or prolonged hospitalization. |
| Reported, severity unclear | Respiratory or pulmonary complication described but severity unclear. |
| Procedure-Related Infection. Infection attributable to the ablation. Includes access-site infection (cellulitis, abscess), endocarditis from catheter-introduced organisms, bacteremia or sepsis from intravascular instrumentation, mediastinitis (may accompany esophageal injury), and post-procedure pneumonia. Cues: fever, elevated white blood cell count, positive blood cultures, wound erythema or drainage, antibiotics for a suspected procedure-related source. EXCLUDES infections unrelated to the procedure. |  |
| Local | Localized access-site infection (cellulitis, superficial wound) managed with oral antibiotics, no hospitalization. |
| Systemic iv antibiotics | Systemic infection (bacteremia, pneumonia, UTI) requiring IV antibiotics or hospitalization, without hemodynamic instability or end-organ damage. |
| Sepsis | Sepsis or septic shock from a procedure-related source (endocarditis, mediastinitis, septic embolism). Requires ICU-level care, vasopressors, or surgical source control. |
| Reported, severity unclear | Infection or related findings described but source, extent, or severity unclear. |
| Procedure-Related Death. Death during or after AF ablation judged related to the procedure or one of its complications. Multifactorial: cardiac tamponade, atrioesophageal fistula, massive stroke, anesthesia event, coronary occlusion, or other procedural causes. Overall incidence ~0.05-0.1%. Narrative may state death outright or describe a course leading to it (refractory cardiac arrest, withdrawal of care). Captures death as the final outcome; the underlying mechanism may also warrant another category (pericardial, stroke, esophageal). |  |
| Intraprocedural | Death during the procedure itself, in the electrophysiology laboratory or operating room. |
| Periprocedural | Death within 30 days, outside the procedural setting, attributed to a procedural complication. |
| Delayed | Death more than 30 days post-ablation attributed to a procedural complication (late atrioesophageal fistula, late PV stenosis with pulmonary consequences). |
| Reported, severity unclear | Death indicated but timing or cause relative to the procedure unclear. |
| Device / Equipment Malfunction. Failure or malfunction of the ablation catheter, energy generator, mapping system, or ancillary equipment (sheaths, transseptal needles, irrigation pump, recording system), regardless of whether it caused patient injury. In MAUDE often filed under the 'Malfunction' event type with Annex A device-problem codes. Cues: catheter fracture, tip detachment, generator error codes, impedance faults, software or firmware failure, irrigation failure, mapping-system crash, inability to deliver energy. If it also caused a clinical complication, co-assign the relevant clinical category. |  |
| No procedure impact | Malfunction identified and resolved without impact; procedure completed with the same or replacement equipment. |
| Procedure altered | Malfunction required a change in strategy, backup equipment, or premature termination, but no direct patient injury. |
| Patient injury | Malfunction directly caused or contributed to a patient injury. Also classify the injury under the appropriate clinical category. |
| Reported, severity unclear | Device malfunction described but procedure or patient impact unclear. |
| No Patient Harm. Report describes a device problem, procedural deviation, near-miss, or other reportable occurrence but explicitly states no patient injury or harm occurred. Accounts for reports filed to satisfy regulatory obligations that do not represent a clinical complication. Cues: 'no patient harm,' 'no adverse outcome,' 'no clinical consequence,' 'no impact to the patient.' Assign only when absence of harm is affirmatively stated, not merely when harm is unmentioned. |  |
| Confirmed no harm | Narrative explicitly and clearly states no patient harm occurred. |
| Probable no harm | Circumstances strongly suggest no harm, but no explicit confirming statement. |
| Reported, severity unclear | Insufficient information to determine whether patient harm occurred. |
| Other Complication. Procedure-related adverse event not fitting any preceding category; a residual for real but uncommon complications. Examples: cardiac valve injury (mitral or tricuspid damage from catheter manipulation, chordal entanglement, new or worsened regurgitation), vasovagal or autonomic response (profound bradycardia, asystole, or hypotension during energy delivery, especially PFA near ganglionated plexi), iatrogenic atrial septal defect from transseptal puncture needing closure, stiff left atrium syndrome, radiation skin injury, contrast allergy or anaphylaxis, anesthesia complications (aspiration, airway injury, drug reaction), cardiogenic shock not from tamponade, acute heart failure, urinary retention, skin burns, musculoskeletal injury from positioning or PFA skeletal-muscle stimulation. Use only when no more specific category fits. |  |
| Minor | Self-limited or minimal intervention; no prolonged hospitalization; resolved without lasting sequelae. |
| Moderate | Requires additional treatment, prolonged hospitalization, or a secondary procedure, without lasting disability or life-threatening consequence. |
| Severe | Life-threatening, lasting disability, major intervention, or significantly altered clinical course. |
| Reported, severity unclear | Complication in this category described but severity insufficient to grade. |

**Table S2:**
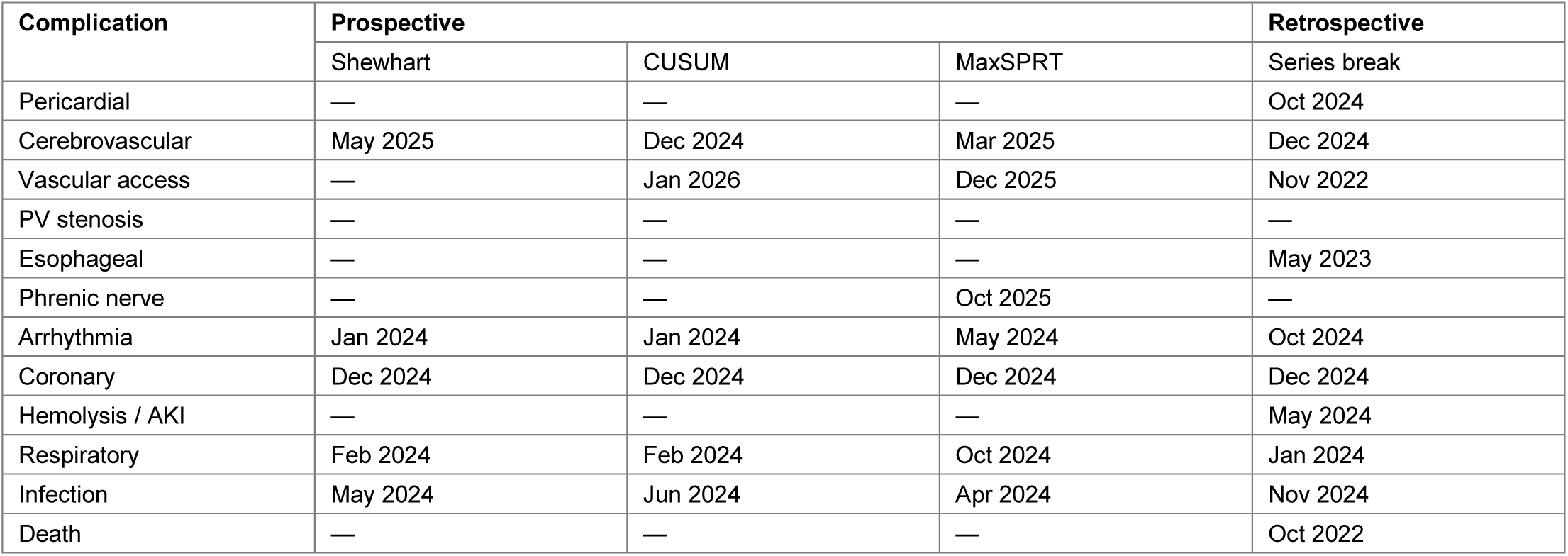
Earliest month that the signal detection methods fired for each complication family. **–** means that method never was triggered (no signal detected). Shewhart, CUSUM, and MaxSPRT are all prospective, and fire when hitting a specific rule. Series break is retrospective, by looking over past window to find a best-fitting timepoint.

## Supplemental Figures

**Figure S1:**
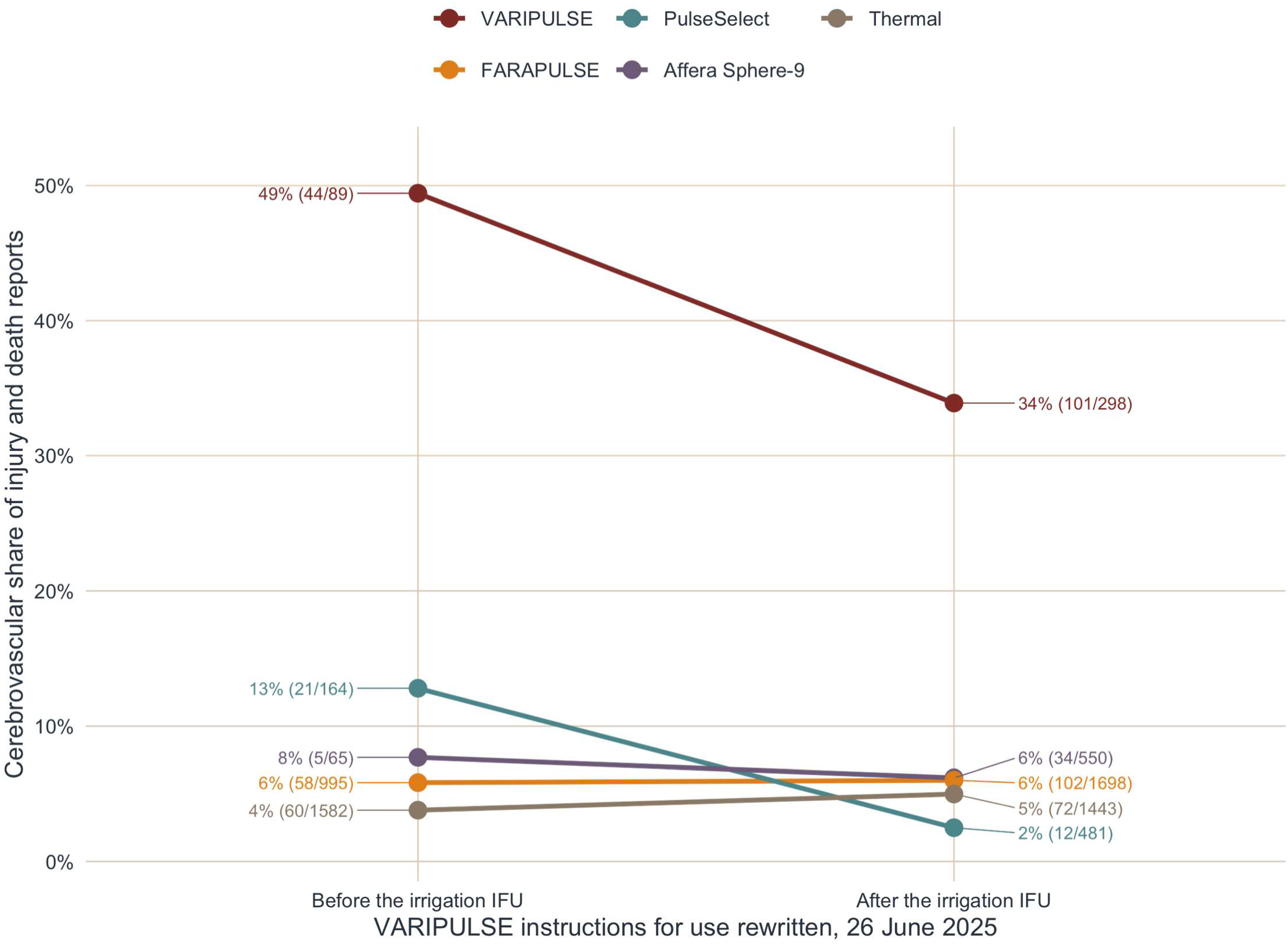
VARIPULSE cerebrovascular reporting before and after the irrigation instructions for use were revised. The VARIPULSE share fell after the revision but remained elevated compared to other platforms. Cerebrovascular share of injury and death reports for each platform and for thermal ablation before and after the VARIPULSE irrigation instructions for use were rewritten on 26 June 2025, with counts at each end. IFU, instructions for use; PFA, pulsed field ablation.

**Figure S2:**
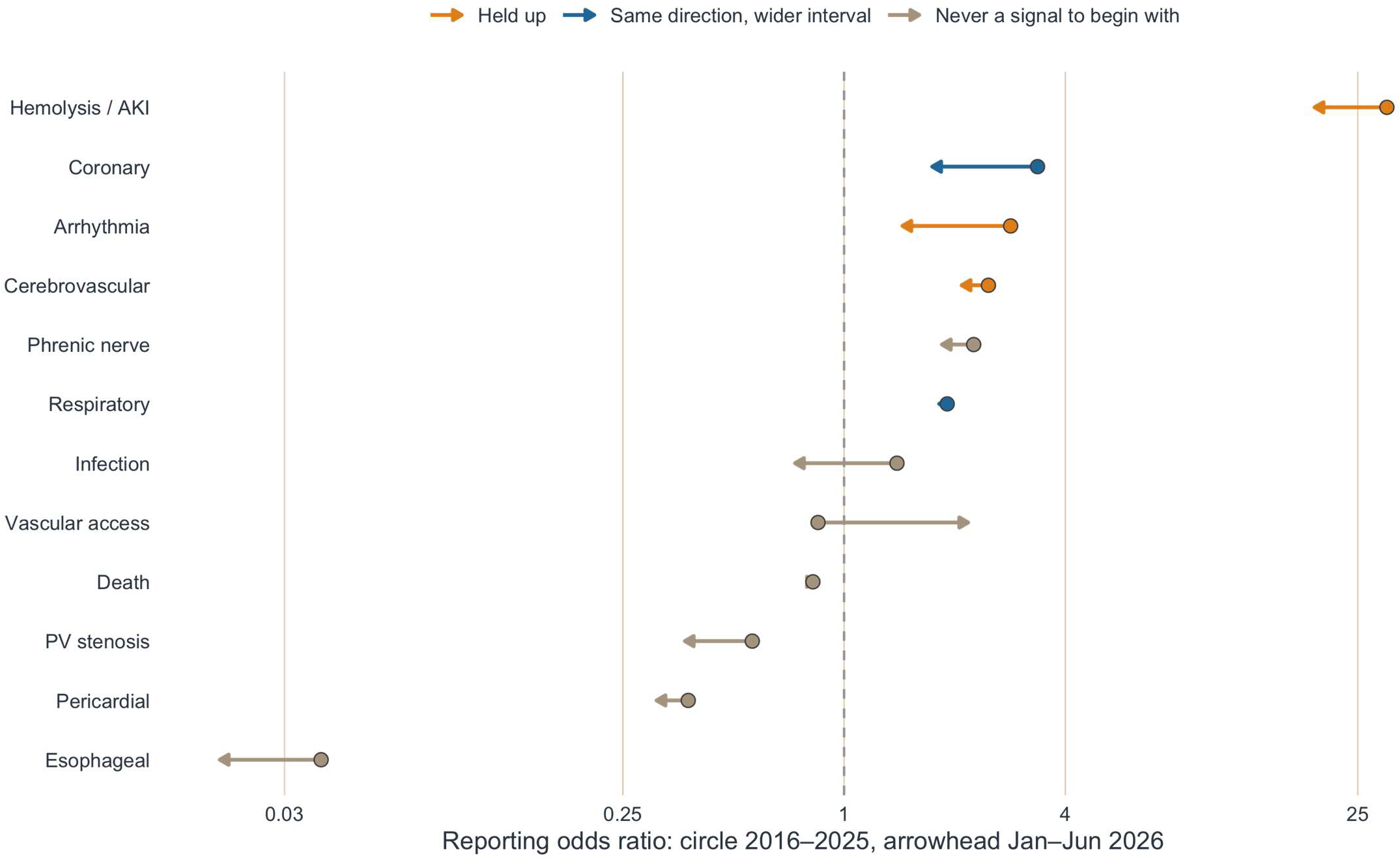
Hold-out re-estimation on reports from January to June 2026. Reporting odds ratio for each complication family estimated on reports received from 2016 through 2025 (circle) and re-estimated on January to June 2026 alone (arrowhead), a period that entered the overall estimates but that no modelling decision was tuned to; colour gives the verdict. AKI, acute kidney injury; PFA, pulsed field ablation; PV, pulmonary vein.

**Figure S3:**
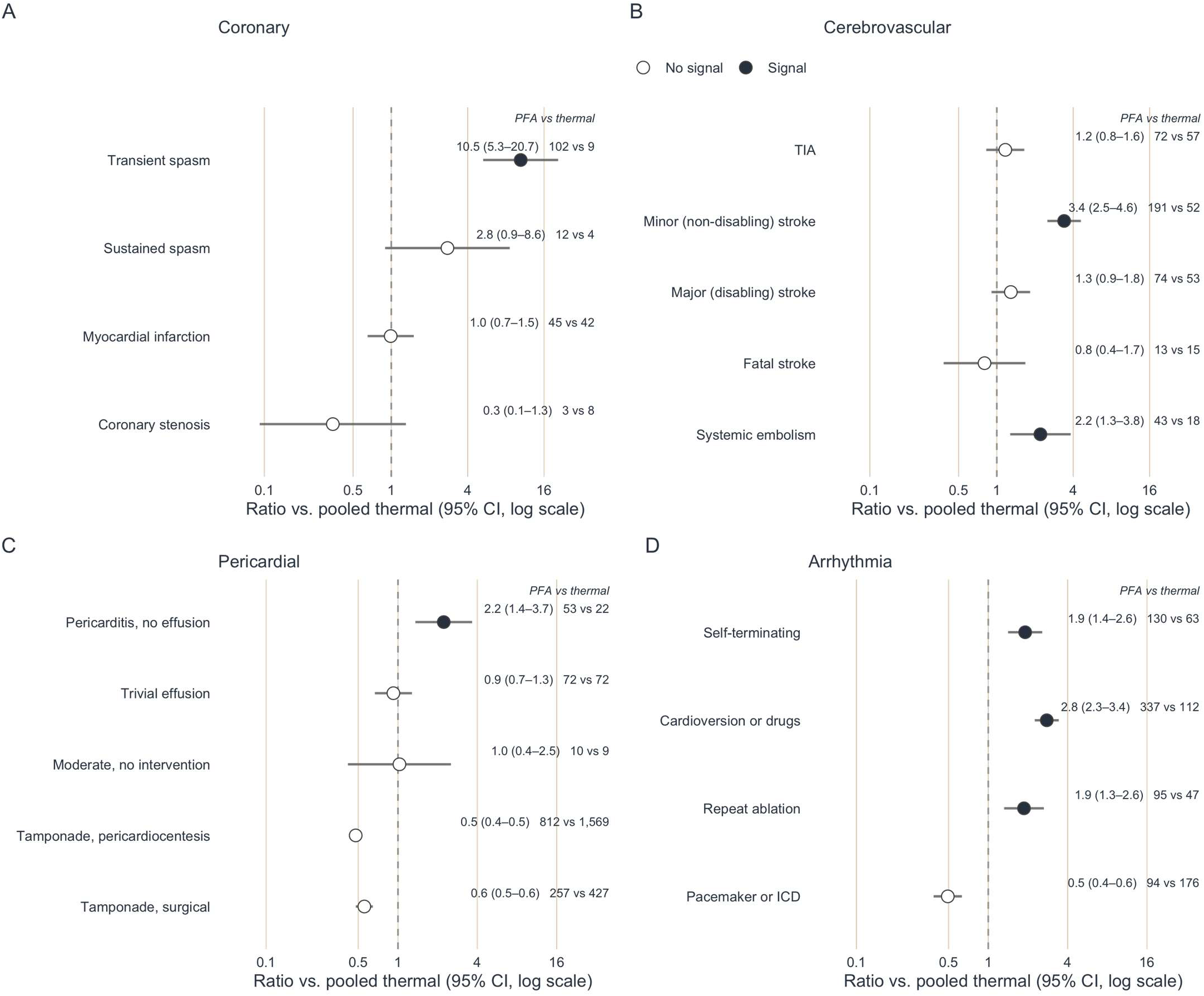
PFA reporting by severity tier within complication families. The coronary excess was confined to transient spasm, every mechanical pericardial tier ran below 1 while pericarditis without effusion ran above it, and the arrhythmia excess did not extend to permanent device implantation. Pulsed field against pooled thermal ablation within each adjudicated severity tier for (A) coronary, (B) cerebrovascular, *(C) pericardial and (D) arrhythmia events, with proportional reporting ratios and 95% confidence intervals on one shared log axis so the four panels can be compared by position; filled points met the signal rule, and the estimate with the pulsed field and thermal counts sits at the right of each row. CI, confidence interval; ICD, implantable cardioverter-defibrillator; PFA, pulsed field ablation; TIA, transient ischemic attack*.

**Figure S4:**
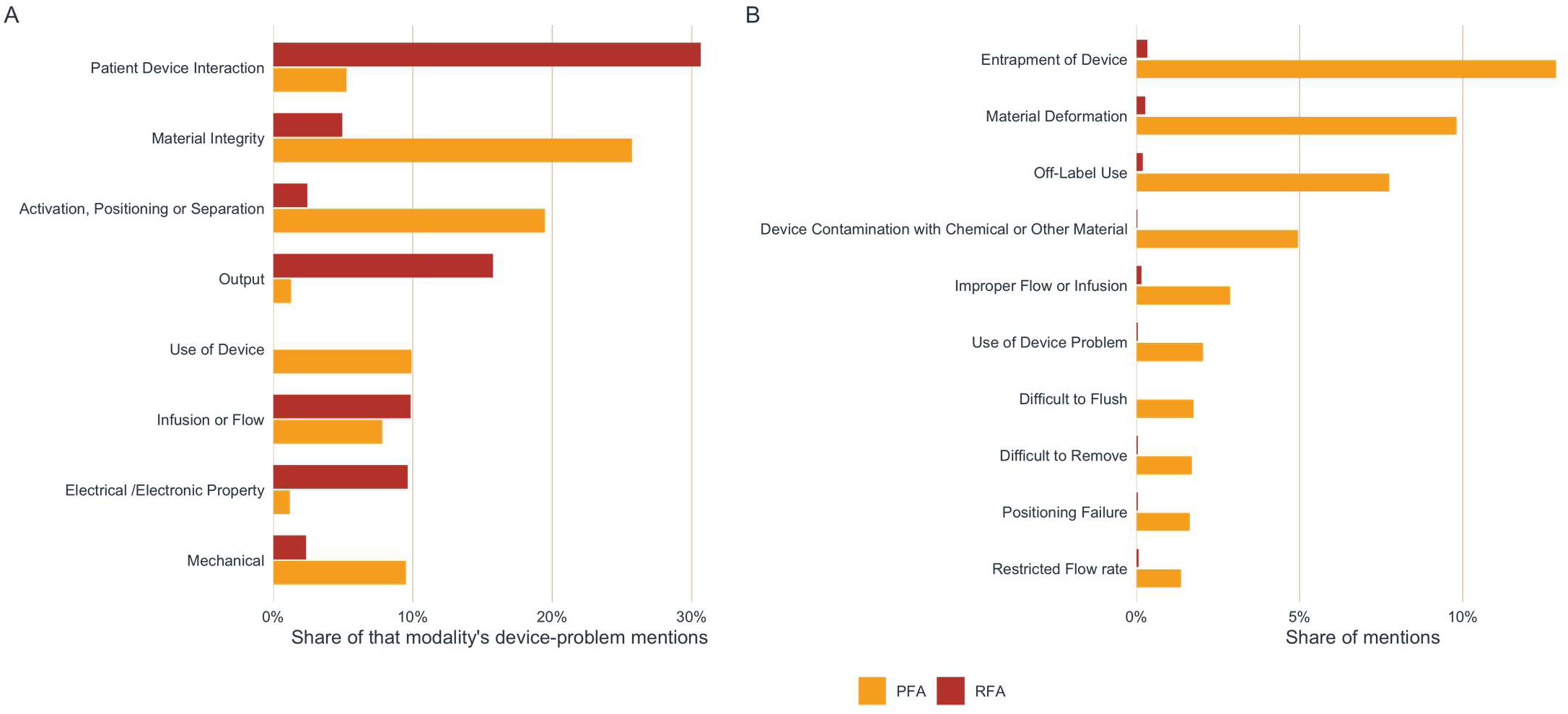
Device problems in malfunction reports. (A) Device-problem families from manufacturer-coded IMDRF Annex A from MAUDE. No overlap with injury/death reports. (B) Most common PFA device issues broken to individual Annex A terms. IMDRF, International Medical Device Regulators Forum; MAUDE, Manufacturer and User Facility Device Experience; PFA, pulsed field ablation.

## Statistical Software

Analyses were performed with R version 4.6.1 (R Core Team, Vienna, Austria) on macOS Golden Gate 27.0,^37^ and the manuscript was rendered with Quarto 1.10.18. MAUDE querying and retrieval from openFDA were through the card package.^38^ The R packages called by the analysis pipeline are listed below with their versions.

| Package | Version | Source |
| --- | --- | --- |
| brand.yml | 0.1.0 | CRAN (R 4.6.0) |
| card | 0.2.0 | local |
| crew | 1.3.2 | CRAN (R 4.6.1) |
| digest | 0.6.39 | CRAN (R 4.6.0) |
| dplyr | 1.2.1 | CRAN (R 4.6.0) |
| ellmer | 0.4.2 | CRAN (R 4.6.1) |
| forcats | 1.0.1 | CRAN (R 4.6.0) |
| ggplot2 | 4.0.3 | CRAN (R 4.6.0) |
| ggraph | 2.2.2 | CRAN (R 4.6.0) |
| ggrepel | 0.9.8 | CRAN (R 4.6.0) |
| ggtext | 0.2.0 | CRAN (R 4.6.1) |
| glue | 1.8.1 | CRAN (R 4.6.0) |
| gt | 1.3.0 | CRAN (R 4.6.0) |
| gtsummary | 2.6.0 | CRAN (R 4.6.1) |
| here | 1.0.2 | CRAN (R 4.6.0) |
| httr | 1.4.9 | CRAN (R 4.6.1) |
| jsonlite | 2.0.0 | CRAN (R 4.6.0) |
| knitr | 1.51 | CRAN (R 4.6.0) |
| lubridate | 1.9.5 | CRAN (R 4.6.0) |
| MASS | 7.3-66 | CRAN (R 4.6.1) |
| openEBGM | 0.10.0 | CRAN (R 4.6.1) |
| patchwork | 1.3.2 | CRAN (R 4.6.0) |
| purrr | 1.2.2 | CRAN (R 4.6.0) |
| readr | 2.2.0 | CRAN (R 4.6.0) |
| rlang | 1.3.0 | CRAN (R 4.6.1) |
| sandwich | 3.1-3 | CRAN (R 4.6.1) |
| scales | 1.4.0 | CRAN (R 4.6.0) |
| sessioninfo | 1.2.4 | CRAN (R 4.6.0) |
| stringr | 1.6.0 | CRAN (R 4.6.0) |
| tarchetypes | 0.14.1 | CRAN (R 4.6.0) |
| targets | 1.12.0 | CRAN (R 4.6.0) |
| tibble | 3.3.1 | CRAN (R 4.6.0) |
| tidygraph | 1.3.1 | CRAN (R 4.6.0) |
| tidyr | 1.3.2 | CRAN (R 4.6.0) |
| tidyselect | 1.2.1 | CRAN (R 4.6.0) |
| tidyverse | 2.0.0 | CRAN (R 4.6.0) |
| withr | 3.0.3 | CRAN (R 4.6.0) |

## REFERENCES

1. Tabaja C, Younis A, Hussein AA, et al. Catheter-Based Electroporation. JACC Clin Electrophysiol. 2023;9:2008–2023.

2. Ekanem E, Reddy VY, Schmidt B, et al. Multi-national survey on the methods, efficacy, and safety on the post-approval clinical use of pulsed field ablation (MANIFEST-PF). EP Eur. 2022;24:1256–1266.

3. Ekanem E, Neuzil P, Reichlin T, et al. Safety of pulsed field ablation in more than 17,000 patients with atrial fibrillation in the MANIFEST-17K study. Nat Med. 2024;30:2020–2029.

4. Schmidt B, Bordignon S, Neven K, et al. EUropean real-world outcomes with Pulsed field ablatiOn in patients with symptomatic atRIAl fibrillation: Lessons from the multi-centre EU-PORIA registry. EP Eur. 2023;25:euad185.

5. Turagam MK, Aryana A, Day JD, et al. Multicenter Study on the Safety of Pulsed Field Ablation in Over 40,000 Patients. JACC. 2026;87:172–193.

6. Reddy VY, Gerstenfeld EP, Natale A, et al. Pulsed Field or Conventional Thermal Ablation for Paroxysmal Atrial Fibrillation. N Engl J Med. 2023;389:1660–1671.

7. Anter E, Mansour M, Nair DG, et al. Dual-energy lattice-tip ablation system for persistent atrial fibrillation: A randomized trial. Nat Med. 2024;30:2303–2310.

8. Shah AS, Zenger B, Wall TS, Bunch TJ. Reappraising CHAMPION AF. Does Placebo Meet Non-Inferiority in Stroke Prevention? Heart Rhythm. 2026;0.

9. Venier S, Vaxelaire N, Jacon P, et al. Severe acute kidney injury related to haemolysis after pulsed field ablation for atrial fibrillation. Europace. 2023;26:euad371.

10. Mohanty S, Casella M, Compagnucci P, et al. Acute Kidney Injury Resulting From Hemoglobinuria After Pulsed-Field Ablation in Atrial Fibrillation. JACC Clin Electrophysiol. 2024;10:709–715.

11. Gianni C, Al-Ahmad A, Elchouemi M, et al. Pulsed Field Ablation–Related Hemolysis: Comparison Between Technologies. Circ Arrhythm Electrophysiol. 2025;18.

12. Masuda H, Nagashima K, Matsumoto N, et al. Lesion area, ablation time, fluoroscopic time, and short-term outcomes among 3 pulsed field ablation systems. Heart Rhythm. 2026;23:1517–1523.

13. Kawamura I, Miyazaki S, Kato R, et al. Comparison of hemolysis with different pulsed field ablation systems. Heart Rhythm. 2026;23:388–395.

14. Reddy VY, Petru J, Funasako M, et al. Coronary Arterial Spasm During Pulsed Field Ablation to Treat Atrial Fibrillation. Circulation. 2022;146:1808–1819.

15. Zhang C, Neuzil P, Petru J, et al. Coronary Artery Spasm During Pulsed Field vs Radiofrequency Catheter Ablation of the Mitral Isthmus. JAMA Cardiol. 2024;9:72.

16. Malyshev Y, Neuzil P, Petru J, et al. Nitroglycerin to Ameliorate Coronary Artery Spasm During Focal Pulsed-Field Ablation for Atrial Fibrillation. JACC Clin Electrophysiol. 2024;10:885–896.

17. Lakkireddy D, Katapadi A, Garg J, et al. NEMESIS-PFA: Investigating Collateral Tissue Injury Associated with Pulsed Field Ablation. JACC Clin Electrophysiol. 2025. Published onlineApril 24, 2025. 10.1016/j.jacep.2025.04.017.

18. Hauser RG. Disproportionate view of pulsed field ablation safety signals. Heart Rhythm. 2026;0.

19. Wazni OM, Chun KRJ, Nair DG, et al. Pulsed Field Ablation as Initial Therapy for Persistent Atrial Fibrillation. N Engl J Med. 2026;394:2407–2418.

20. Hammadeh BM, Aldib L, Challa A, Gerlt D, Al Mubaid A, Kohli U. Real-World Adverse Events of Pulsed Field Ablation for Atrial Fibrillation: A Comprehensive Analysis of the FDA MAUDE Database. Heart Rhythm. 2026. Published onlineJuly 31, 2026. 10.1016/j.hrthm.2026.07.032.

21. Ferro EG, d’Avila A, Reynolds MJ, et al. Risk of Stroke and TIA With Pulsed Field Compared With Radiofrequency Ablation for Atrial Fibrillation. Circulation. 2026. Published onlineAugust 25, 2026. 10.1161/CIRCULATIONAHA.126.081092.

22. Castrejón-Castrejón S, Heeger C-H, Veltmann C, et al. Stroke and systemic embolism following atrial fibrillation ablation: The EMBOL-AF Global Registry. Eur Heart J. 2026:ehag217.

23. Duggirala HJ, Herz ND, Caños DA, et al. Disproportionality analysis for signal detection of implantable cardioverter–defibrillator-related adverse events in the Food and Drug Administration Medical Device Reporting System. Pharmacoepidemiol Drug Saf. 2012;21:87–93.

24. Shah AS, Engels M, Freedman RA, et al. Real-World Safety Profile of the Extravascular (EV) Implantable Defibrillator System: Insights From the MAUDE Database. Circ Arrhythm Electrophysiol. 2025;18:e013831.

25. Tzeis S, Gerstenfeld EP, Kalman J, et al. 2024 European Heart Rhythm Association/Heart Rhythm Society/Asia Pacific Heart Rhythm Society/Latin American Heart Rhythm Society expert consensus statement on catheter and surgical ablation of atrial fibrillation. Eur Eur Pacing Arrhythm Card Electrophysiol J Work Groups Card Pacing Arrhythm Card Cell Electrophysiol Eur Soc Cardiol. 2024;26:euae043.

26. Joglar JA, Chung MK, Armbruster AL, et al. 2023 ACC/AHA/ACCP/HRS Guideline for the Diagnosis and Management of Atrial Fibrillation: A Report of the American College of Cardiology/American Heart Association Joint Committee on Clinical Practice Guidelines. Circulation. 2024;149:e1–e156.

27. Evans SJW, Waller PC, Davis S. Use of proportional reporting ratios (PRRs) for signal generation from spontaneous adverse drug reaction reports. Pharmacoepidemiol Drug Saf. 2001;10:483–486.

28. Haldane JBS. The mean and variance of chi-square, when used as a test of homogeneity, when expectations are small. Biometrika. 1940;31:346–355.

29. Anscombe FJ. On estimating binomial response relations. Biometrika. 1956;43:461–464.

30. Bate A, Lindquist M, Edwards IR, et al. A Bayesian neural network method for adverse drug reaction signal generation. Eur J Clin Pharmacol. 1998;54:315–321.

31. Dumouchel W. Bayesian Data Mining in Large Frequency Tables, with an Application to the FDA Spontaneous Reporting System. Am Stat. 1999;53:177–190.

32. Benjamini Y, Hochberg Y. Controlling the False Discovery Rate: A Practical and Powerful Approach to Multiple Testing. J R Stat Soc Ser B Stat Methodol. 1995;57:289–300.

33. Bernal JL, Cummins S, Gasparrini A. Interrupted time series regression for the evaluation of public health interventions: A tutorial. Int J Epidemiol. 2016:dyw098.

34. Laney DB. Improved Control Charts for Attributes. Qual Eng. 2002;14:531–537.

35. Page ES. Continuous Inspection Schemes. Biometrika. 1954;41:100–115.

36. Kulldorff M, Davis RL, Kolczak† M, Lewis E, Lieu T, Platt R. A Maximized Sequential Probability Ratio Test for Drug and Vaccine Safety Surveillance. Seq Anal. 2011;30:58–78.

37. R Core Team. R: A language and environment for statistical computing. Vienna, Austria: R Foundation for Statistical Computing, 2026.

38. Shah AS. Card: Cardiovascular Applications in Research Data. 2024.

39. Cho MS, Lee S-R, Black-Maier E, et al. Complications associated with pulsed field ablation vs radiofrequency catheter ablation of atrial fibrillation. Heart Rhythm. 2024:S1547527124035288.

40. Shoura SJ, Singireddy S, Coombes K, et al. Real-World Safety of Two Pulsed Field Ablation Catheters for Atrial Fibrillation: A MAUDE Analysis. Pacing Clin Electrophysiol. 2026:pace.70396.

41. Isenegger C, Arnet R, Jordan F, et al. Incidence of acute pericarditis after pulsed-field ablation for the treatment of atrial fibrillation. Europace. 2024;26:euae180.

42. Inaba O, Miyazaki S, Nitta J, et al. Comparative Analysis of Safety Profiles of Pulsed Field and Thermal Energy Atrial Fibrillation Ablation: Insight From the Multicenter Real-World Ablation Registry. J Cardiovasc Electrophysiol. 2026:jce.70474.

43. Porterfield C, Dominic P, Khaykin Y, et al. Variable loop circular catheter pulsed field ablation in real-world practice: Low complication rates across patient and procedural characteristics in the REAL AF registry. Heart Rhythm. 2026;23:1892–1898.

44. Freeman JV, Desai NR, Tan Z, et al. Atrial fibrillation ablation energy source transition and adjunctive lesion set utilization after commercial release of pulse field ablation. Heart Rhythm. 2026:S1547527126024975.

